# TNFRSF13B genotype governs susceptibility to ARDS through IgG-mediated complement activation

**DOI:** 10.64898/2026.06.02.26354763

**Authors:** Lwar Naing, Mayara G de Mattos Barbosa, Ian P Connell, Jeffrey Chicca, Ziyin Zhao, Nerissa A Reister, Anna Bruchez, Neil S. Greenspan, Grace A. McComsey, Jeffrey L Platt, Marilia Cascalho

## Abstract

Acute respiratory distress syndrome (ARDS) is a devastating complication of respiratory infections; however, the biological mechanisms that initiate its onset are poorly defined. Here we show that *TNFRSF13B* polymorphisms increase the risk of ARDS following SARS-CoV-2 infection up to 7.4-fold compared to the WT genotype. The increased risk was not due to immune-deficiency or impaired virus neutralization. On the contrary, *TNFRSF13B* mutant subjects mounted better antibody neutralization compared to subjects with WT *TNFRSF13B*. However, IgG from subjects expressing TNFRSF13B variants had less sialic acid, terminal galactose, and fucose than IgG from subjects with a WT genotype. Moreover, IgG from *TNFRSF13B* mutant subjects exhibited increased recruitment of complement factors. Thus, besides well-known actions governing plasma cell differentiation, *TNFRSF13B* impacts both affinity maturation and effector functions of IgG in ways that independently govern complement activation controlling inflammatory responses known to trigger ARDS.

**Summary:** Acute respiratory distress syndrome (ARDS) is a severe complication of respiratory infections. We show that common TNFRSF13B polymorphisms increase ARDS risk after SARS-CoV-2 infection by up to 7.4-fold by altering IgG glycosylation, enhancing complement activation, and amplifying inflammatory pathways despite preserved or improved viral neutralization.

## Introduction

Acute respiratory distress syndrome (ARDS) is a clinical syndrome defined by rapid-onset respiratory failure, diffuse alveolar damage, and severe hypoxemia resulting from a wide range of infectious and noninfectious insults (Matthay et al., 2019; Williams et al., 2021). ARDS reflects breakdown of the alveolar-capillary barrier (Gonzales et al., 2015), driven by dysregulated innate immune activation (Zhang et al., 2025), endothelial and epithelial injury (Vassiliou et al., 2020), and alterations in pulmonary vascular permeability (Gonzales et al., 2015). Unexplained is why only a subset of exposed individuals progress to fulminant lung injury and what early biological events irreversibly trigger ARDS, a lethal complication of SARS-CoV-2 infection.

The *Tumor Necrosis Factor Receptor Superfamily Member 13B* (*TNFRSF13B*), encodes the Transmembrane Activator and CAML Interactor (TACI), a receptor that promotes B cell differentiation and antibody production (He et al., 2010; Mantchev et al., 2007). *TNFRSF13B* is highly polymorphic, with over 743 missense and 304 synonymous mutations according to Ensembl database (Cascalho and Platt, 2021). Although genes associated with immune recognition and related processes can be highly polymorphic (e.g. HLA), the nature of *TNFRSF13B* polymorphisms is quite unique. Unlike polymorphic genes essential for immune functions *TNFRSF13B* has an excess of single nucleotide variants predicted to cause loss of function (36 observed versus 26 predicted) based on the calculated mutation rate for the locus by GnomAD browser (Chen et al., 2024). Furthermore, a subset of *TNFRSF13B* mutations have been associated with common variable immunodeficiency (CVID) (Freiberger et al., 2012), primary IgA deficiency (Salzer et al., 2005) or tonsillar hypertrophy (Speletas et al., 2013). However, most individuals with *TNFRSF13B* mutations are healthy (Abolhassani et al., 2025; Cascalho and Platt, 2021). Moreover, common TNFRSF13B variants such as P251L are observed at frequencies of approximately 11% in European populations (Chen et al., 2024). These findings led us to hypothesize that *TNFRSF13B* polymorphisms may provide a selective advantage to the host at a cost that might include increased risk for immunodeficiency (Platt et al., 2021), autoimmunity (Barroeta Seijas et al., 2012) and inflammatory responses (de Mattos Barbosa et al., 2021a), thereby explaining the persistence of many *TNFRSF13B* mutations in the population.

Given that SARS-CoV-2 emerged as a novel pathogen to which the global population was immunologically naïve, we hypothesized that genetic diversity of *TNFRSF13B* could determine host susceptibility or resistance to severe disease, including ARDS. While some TNFRSF13B specific variants might increase susceptibility to severe disease, including progression to ARDS, others might confer protection, thereby being more prevalent in individuals who experienced milder illness. To test the hypothesis, we performed targeted sequencing of all five coding exons of *TNFRSF13B* in a cohort of 108 patients with confirmed SARS-CoV-2 infection, 66 of which met criteria for severe disease. Understanding how *TNFRSF13B* polymorphisms determine immune responses to a new pathogen like SARS-CoV-2 illuminates broader principles of host-pathogen interactions and genetic determinants of disease severity.

## Results

### TNFRSF13B variants are associated with severe SARS-CoV-2 infection

To investigate whether and how TNFRSF13B variants might impact Coronavirus disease 2019 (COVID-19) pathogenesis, we sequenced the five exons of *TNFRSF13B* in 108 subjects diagnosed with SARS-CoV-2 infection. Of 108 subjects with SARS-CoV-2, 40 (37%) had at least one allele encoding *TNFRSF13B* missense mutations (Figure 1A). The missense mutations included one C104R and one P116T variant in exon 3, and one G190R variant in exon 4, each identified in a single subject (0.9%). Five subjects had mutations encoding the V220A variant (4.6%). Twenty-four subjects had mutations encoding monoallelic P251L (22.2%) and 8 subjects were homozygous for the P251L variant (7.4%).

**Figure 1:**
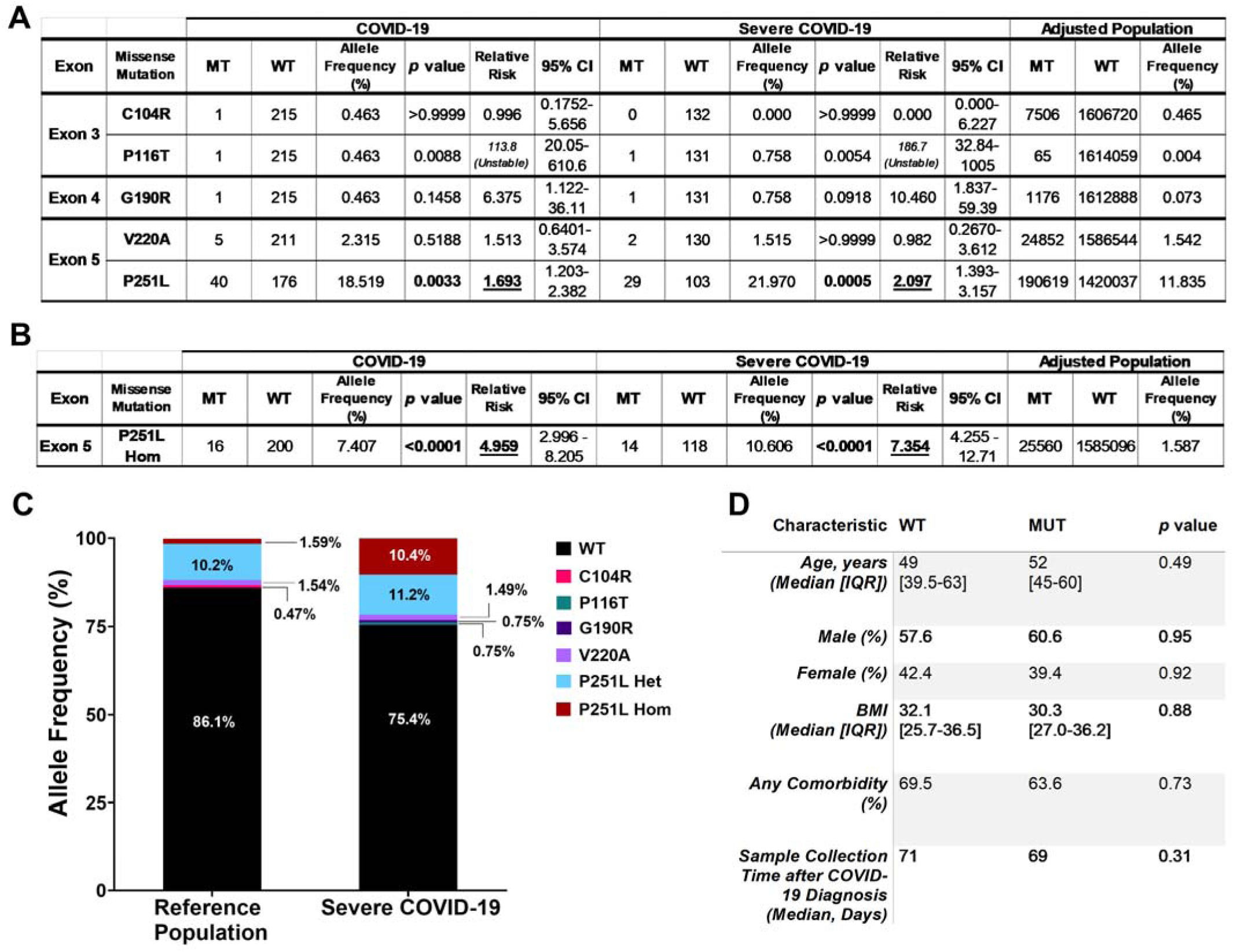
*TNFRSF13B* missense mutation frequencies in subjects with SARS-CoV-2 infection. *TNFRSF13B* exons 1 to 5 were amplified by PCR and sequenced by the Sanger method. **A.** Frequency of missense mutations is shown in the study group and as reported by the GnomAD 4.1 browser. GnomAD allele frequencies were adjusted by computing a weighted allele frequency to reflect the demographics of the study population. Significant *p* values identified by Fisher’s exact test are underlined and the corresponding Relative Risk values are underlined and bolded. “Instability” is indicated if confidence intervals are large due to small sample size. **B.** The frequencies of P251L alleles in homozygous subjects (P251L Hom) are indicated separately. **C.** Allele frequencies of reference population and severe COVID-19 group are also shown. **D.** Multivariable analysis of COVID-19 subjects with or without TNFRSF13B variants. Comorbidities included hypertension, diabetes, heart disease, chronic lung disease, kidney disease, and cancer. Continuous variables are presented as median [interquartile range (IQR)] and were compared using the Wilcoxon rank-sum test. Categorical variables such as age, sex, and comorbidities were compared using the chi-square test. Sample collection time after diagnosis was analyzed using Mann-Whitney Test.

Of 108 subjects diagnosed with SARS-CoV-2 infection during their hospital or the medical center visit, 66 met the criteria for severe SARS-CoV-2 infection and developed ARDS. Severe COVID-19 was defined primarily based on clinical documentation in the medical record, including diagnoses of ARDS, ICU admission, and/or requirement for mechanical ventilation or ECMO (32/66 cases). For cases lacking explicit diagnostic descriptions, intubation status was used to classify severity (20/66 cases). In remaining cases (14/66 cases), physiologic criteria, including SpO < 80%, respiratory rate > 25 breaths per minute, or oxygen flow rate > 2 L/min, as well as a pneumonia diagnosis were used to assign patients to the severe group. Patients’ clinical and demographic information is provided in Extended Data Table 1.

Among the 66 subjects with severe SARS-CoV-2 infection, 26 (39.4%) carried at least one *TNFRSF13B* mutated allele. The mutated alleles included one subject with a P116T allele (1.5%), one subject with a G190R allele (1.5%), two subjects with one V220A allele each (3%). Twenty-two subjects were P251L carriers (33.3%), 7 individuals were homozygous (10.6%) and 15 (22.7%) were heterozygous (Figure 1A). *TNFRSF13B* allele frequencies between the reference population and severe group is also shown (Figure 1C).

Based on allele frequencies in the Genome Aggregation Database (gnomAD browser v4.1.0) (Chen et al., 2024), expression of biallelic *TNFRSF13B* P251L increased the risk for severe SARS-CoV-2 infection by 7-fold (p <0.0001, Figure 1B) and in those of Asian ethnicity by 21-fold (p <0.0001, Extended Data Table 2). Multivariate analysis revealed that the association of TNFRSF13B variants and severe disease was not due to differences in age, sex, BMI, or comorbidities as those variables were comparable in subjects with WT or TNFRSF13B variants (Figure 1D). The findings support the central premise that *TNFRSF13B* polymorphisms increase susceptibility to severe respiratory failure following viral infections.

### Subjects with TNFRSF13B variants do not have antibody deficiency

Since TNFRSF13B controls antibody production (de Mattos Barbosa et al., 2021a; Tsuji et al., 2011; Tsuji et al., 2014), we asked what changes in antibody responses in subjects with *TNFRSF13B* mutations could contribute to disease severity. Subjects with *TNFRSF13B* missense mutations and SARS-CoV-2 infection did not exhibit Ig-deficiency (Extended Data Fig. 1, Extended Data Fig. 2). Furthermore, severity of SARS-CoV-2 pulmonary disease in subjects with *TNFRSF13B* mutations was evidently not a consequence of hypogammaglobulinemia. In fact, those with missense mutations and severe SARS-CoV-2 infection had increased serum IgM, IgG, and IgA concentrations (2.1-fold, 1.3-fold, and 2.7-fold, respectively) compared to values measured in subjects with WT *TNFRSF13B* (Figure 2A-C). Similarly, when the analysis was restricted to subjects with the P251L variant, we did not see evidence of Ig deficiency compared to WT subjects (Extended Data Fig. 2). Thus, although *TNFRSF13B* mutations have been sporadically associated with hypogammaglobulinemia (Salzer et al., 2009) (Pulvirenti et al., 2016), the subjects with *TNFRSF13B* mutations and severe disease had increased rather than decreased Ig in blood.

**Figure 2:**
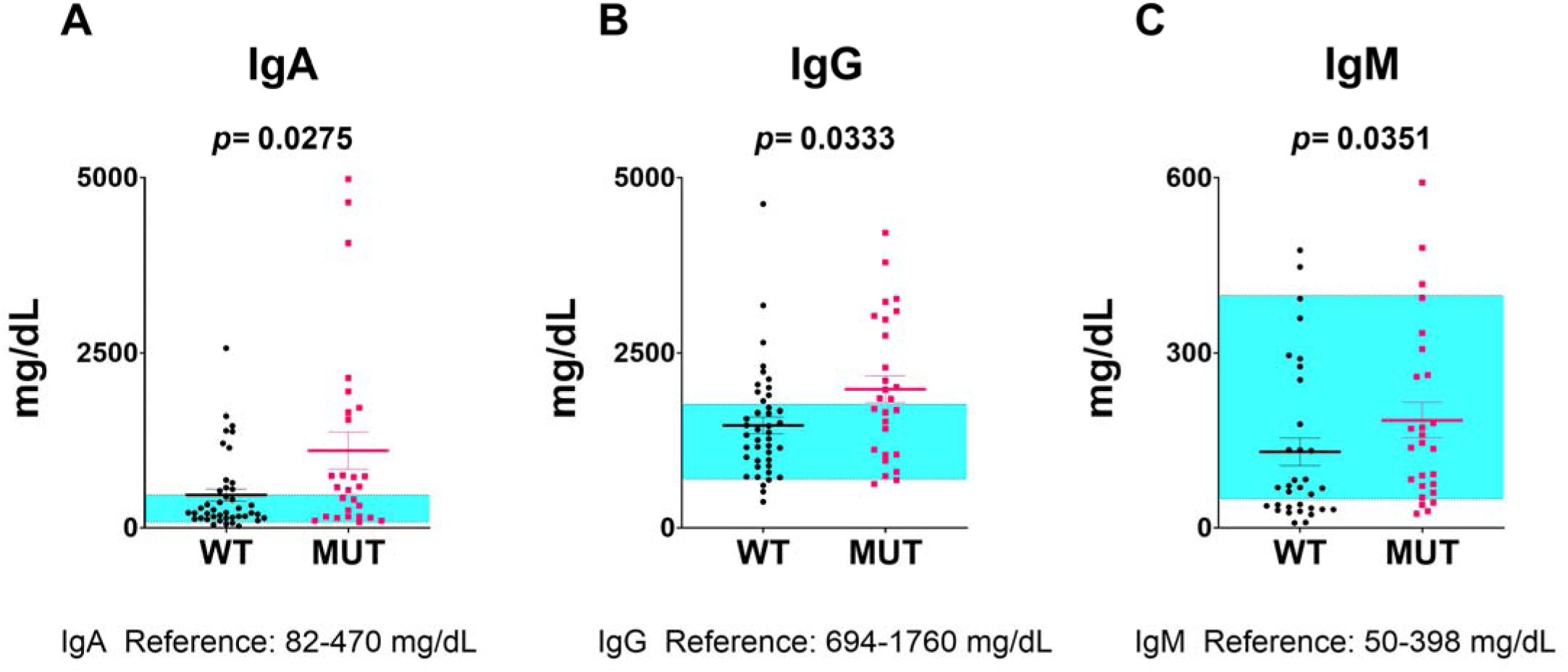
Plasma immunoglobulin concentrations in subjects diagnosed with severe SARS-CoV-2 infection. **A.** Serum IgA, **B.** IgG, and **C.** IgM measured as mg/dL in TNFRSF13B variant carriers (MUT) compared to subjects with WT alleles (WT, n = 42; MUT, n = 23 for IgA and IgG; WT, n = 36; MUT, n = 22 for IgM). The blue bar depicts the normal range (Gonzalez-Quintela et al., 2008). Groups were compared by a two-tailed Mann-Whitney Test in GraphPad Prism. *p* values are indicated on the graphs.

### Mutations of *TNFRSF13B* do not impair Wuhan-Hu-1 SARS-CoV-2 plasma neutralization

Given the importance of *TNFRSF13B* in maturation of B cells and plasma cells, one might think mutations undermining the function of the receptor might impair generation of virus-neutralizing antibodies. Yet, *TNFRSF13B* missense mutations had no appreciable impact on development of antibodies that neutralize SARS-CoV-2. Subjects with *TNFRSF13B* missense mutations had similar concentrations of anti-SARS-CoV-2 spike protein S1/S2 IgG antibodies compared to subjects with WT *TNFRSF13B* (Figure 3A). To determine if defective SARS-CoV-2 neutralization contributed to severe disease in subjects with *TNFRSF13B* missense mutations, we tested SARS-CoV-2 neutralization using a luciferase reporter lentivirus pseudotyped with either the Wuhan-Hu-1 SARS-CoV-2Δ19AA spike protein or the Delta variant spike (de Mattos Barbosa et al., 2021b). Luciferase-based neutralization assays using target cells transduced with SARS-CoV-2 spike-pseudotyped lentiviral particles are commonly used and highly sensitive (Cruz-Cardenas et al., 2022). Plasma from *TNFRSF13B* variant carriers neutralized SARS-CoV-2 (Wuhan-Hu-1 strain) better by 5-fold, on average, than the plasma from subjects with WT *TNFRSF13B*. (ID_50_ *p=* 0.0627 and ID_80_ *p=* 0.0793, respectively) (Figure 3B). These results indicate that deficient virus neutralization is not likely to contribute to the increased propensity of subjects with *TNFRSF13B* common mutations to develop severe disease.

**Figure 3:**
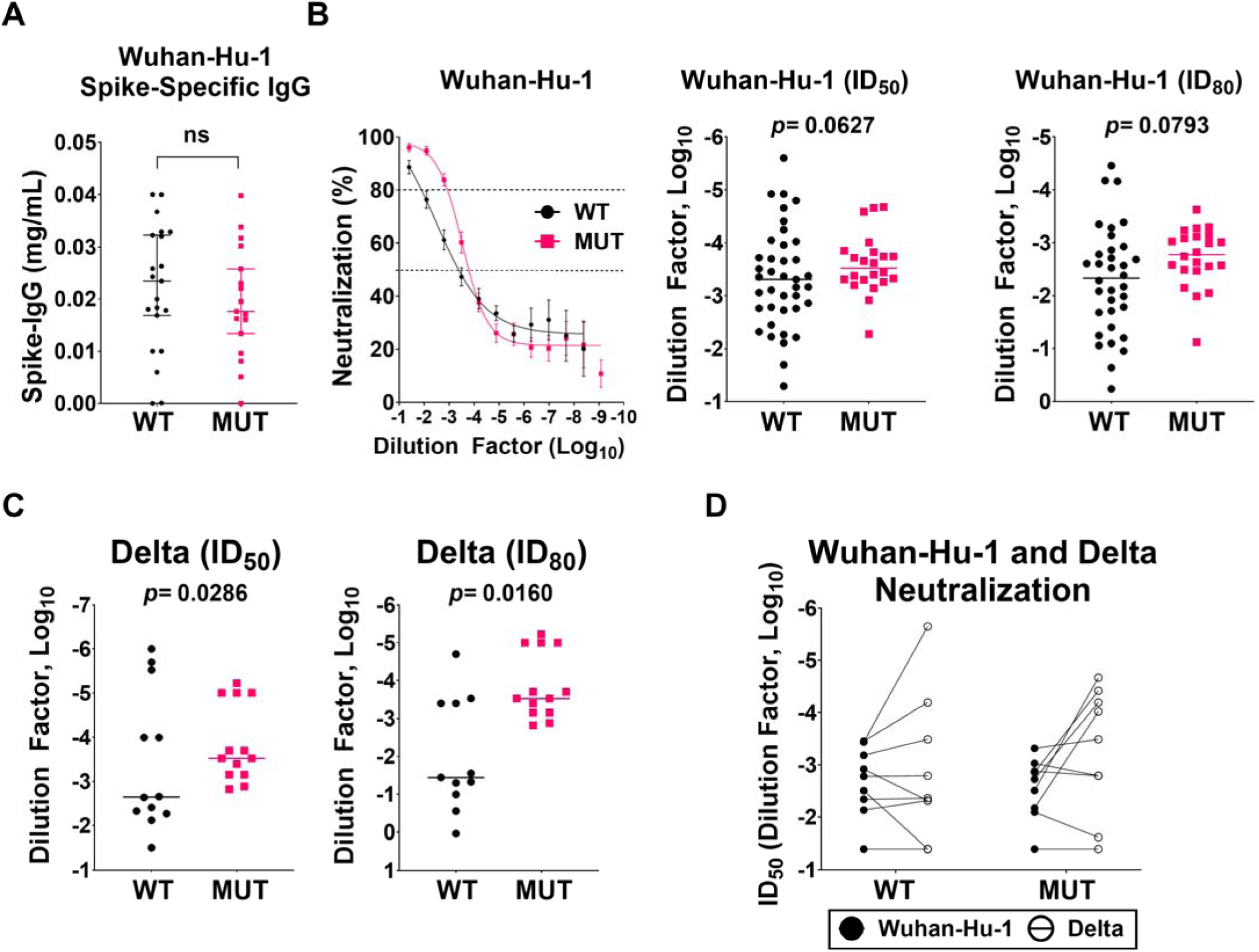
Anti-spike IgG concentrations and virus neutralization by plasma antibodies of SARS-CoV-2 infected individuals with or without TNFRSF13B variants. **A.** Anti-spike IgG was measured in the plasma by ELISA using a commercially purchased Wuhan-Hu-1 spike protein S1/S2 (WT, n= 21; MUT, n= 17). **B and C.** Neutralization of antibodies was measured using Wuhan-Hu-1 spike pseudotyped viruses (WT, n = 39; MUT, n= 22) or Delta spike pseudotyped viruses (WT, n = 12; MUT, n= 13). Scatter plots display individual ID_50_ and ID_80_ values. **D.** Paired analysis of plasma neutralization from the same patients against Wuhan-Hu-1 and Delta spike pseudotyped viruses depicted as ID_50_ (WT, n= 8; MUT, n= 8). Comparisons of anti-spike IgG concentrations were examined using Welch’s t test. *p* values for ID_50_ and ID_80_ comparisons were determined using Kolomogorov-Smirnov test in GraphPad Prism (v.10).

To investigate how *TNFRSF13B* and its variants thereof impact the breadth of neutralization of anti-SARS-CoV-2 antibodies, we asked whether antibodies generated before emergence of the Delta strain would neutralize that viral strain, as the plasma samples were collected during early pandemic and prior to vaccine availability. In a neutralization assay using Delta pseudotyped viruses, plasma from subjects with *TNFRSF13B* variants neutralized the SARS-CoV-2 Delta pseudotyped viruses more efficiently than plasma from WT subjects (ID_50_ *p=* 0.0286, and ID_80_ *p=* 0.016) (Figure 3C). The fraction of subjects exhibiting broad neutralization was increased among subjects with *TNFRSF13B* mutations (44%) relative to the fraction of subjects with WT sequences (22%) (Figure 3D). Thus, mutations of *TNFRSF13B* do not impair and might even improve protective antibody responses to diversifying viruses.

### Subjects with TNFRSF13B variants produce spike-specific antibodies with increased VH somatic hypermutation and more IgG1 and IgG3 compared to subjects with WT TNFRSF13B

We next sought to delineate what antibody properties might contribute to the enhanced neutralization observed in subjects with *TNFRSF13B* mutations. To investigate, we isolated anti-spike-specific B cells from 11 individuals diagnosed with SARS-CoV-2 infection: 5 subjects carrying *TNFRSF13B* missense mutations (1 subject with heterozygous C104R, 1 subject with heterozygous G190R, 2 subjects with heterozygous P251L, and 1 subject with homozygous P251L) and 6 subjects with WT alleles (de Mattos Barbosa et al., 2021b). Monoclonal antibodies were generated from B cells isolated by panning on recombinant S1-C-6HIS-Avi spike protein (Abclonal) immobilized via rabbit anti-Avi-tag antibodies. Single cell Ig H+L sequences were obtained by Next GEM Single Cell V(D)J sequencing followed by NGS. Data were processed as described in (Alamyar, 2012) (Giudicelli et al., 2005) and annotated with IgBLAST (Ye et al., 2013) using default parameters and The International Immunogenetics Information System, IMGT (Alamyar, 2012; Giudicelli et al., 2005; Rogosch et al., 2012). The resulting data were imported into ImmuneDB (Rosenfeld et al., 2018b) for further downstream analyses.

We identified 2473 Ig heavy chain (HC) clonotypes in WT individuals and 1293 HC clonotypes in individuals with mutant alleles. Next, monoclonal antibodies (mAbs; heavy chain + light chain) selected from IgG and IgA HC sequences with the highest frequency of somatic mutations were produced by the University of Michigan Vector core according to established protocols (de Mattos Barbosa et al., 2021b). Forty-three antibodies were produced as IgG1. We obtained 22 mAbs from 3 subjects expressing the WT *TNFRSF13B* gene and 21 mAbs from 2 subjects expressing G190R and C104R heterozygous *TNFRSF13B* variants.

Isotype profiling of the IgH repertoires revealed that *TNFRSF13B* mutation carriers exhibited a distinct immunoglobulin isotype distribution compared to WT controls. Specifically, carriers of *TNFRSF13B* missense mutations produced significantly more IgG1, IgG3, and IgA1, accompanied by a 20% reduction in IgM levels (Figure 4A). Given that IgG1 and IgG3 are the most potent activators of the classical complement pathway via high-affinity binding to C1q (Frischauf et al., 2024; Richardson et al., 2019) these findings suggested that *TNFRSF13B* variants may promote heightened complement activation by antibodies.

**Figure 4:**
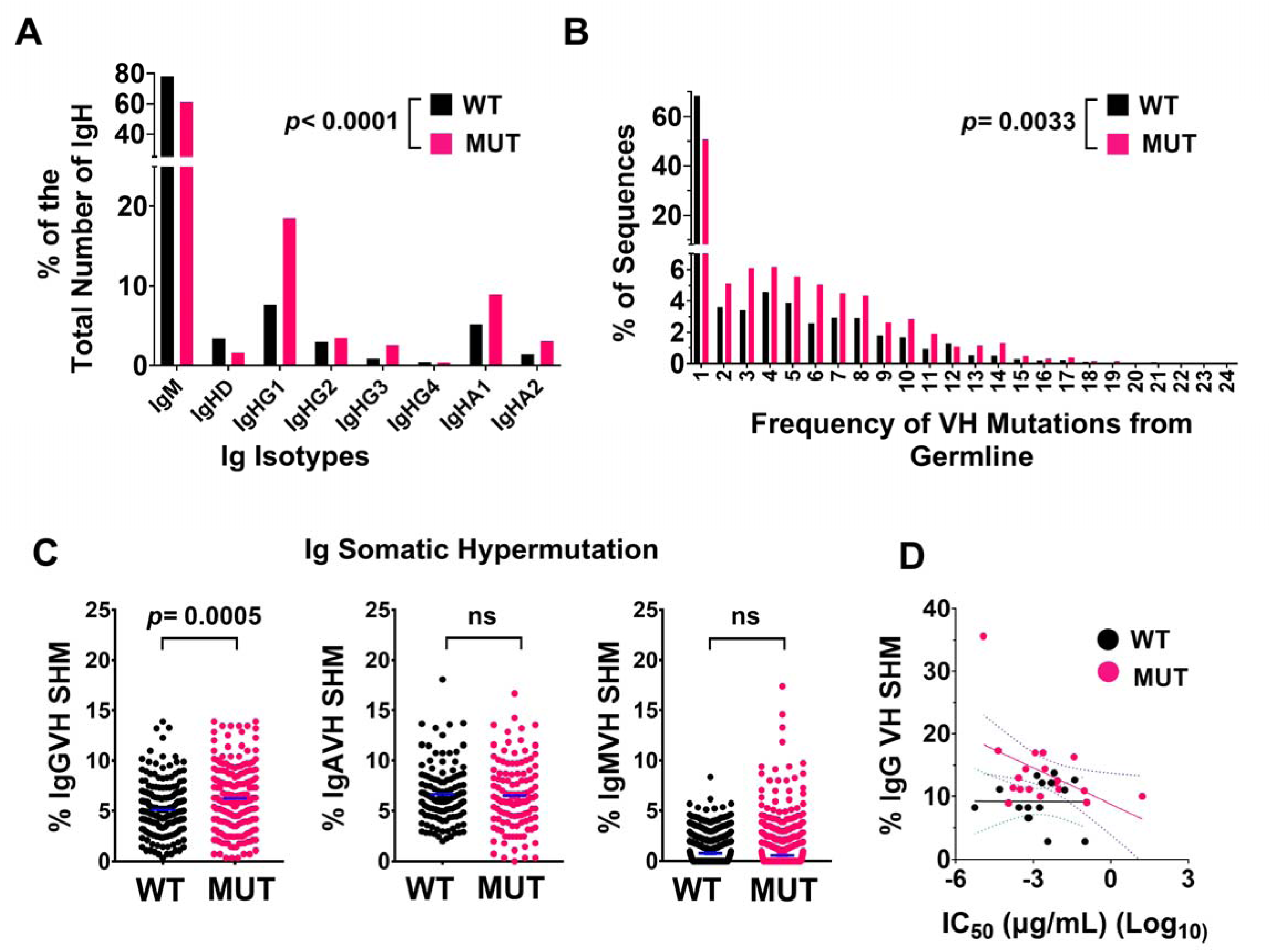
*TNFRSF13B* mutant alleles increase frequency of IgG1, IgG3, IgA1, and VH somatic hypermutation of IgG. **A-C,** Single-cell Ig sequences were obtained from Wuhan-1 Spike–specific memory B cells isolated from 11 subjects (6 subjects with *TNFRSF13B* WT alleles, 1 subject with two P251L alleles, 2 subjects with monoallelic P251L allele, 1 subject with monoallelic G190R and 1 subject with monoallelic C104R). A total of 2,473 Ig heavy-chain (HC) clonotypes were analyzed from WT subjects and 1,293 from mutant (MUT) subjects. **A.** Frequency (%) of each IgH isotype among productive HC sequences. Chi-square analysis revealed significant differences between WT and mutant groups (*p* < 0.0001). **B.** Histogram of VH mutation frequencies. Paired analysis of distributions showed significant differences (*p*= 0.0033). **C.** VH somatic hypermutation (distance from closest germline, NCBI IgBlast) for IgM, IgG, and IgA clonotypes. Only IgG sequences differed significantly between groups (*p* = 0.0005, Mann-Whitney test). **D.** 22 mAbs from 3 WT subjects and 21 mAbs from 2 subjects with mutant alleles (G190R, C104R) were generated. All antibodies were IgG1 and tested for neutralization of Wuhan-Hu-1 spike pseudotyped viruses. Graph shows VH mutation frequency vs IC_50_, µg/mL(Log_10_). Linear regression revealed a negative correlation between VH mutation frequency and IC50 for mutant-derived antibodies (*p*= 0.04), but not for WT-derived antibodies.

Furthermore, analysis of somatic hypermutation frequencies in the variable heavy (VH) regions showed that spike-specific IgG sequences from *TNFRSF13B* variant carriers had a significantly higher average mutation frequency (6.2%) compared to those from WT subjects (4.8%) (Figure 4B). This increase was confined to IgG sequences, with no significant differences observed in IgM or IgA (Figure 4C). To assess functional consequences, we evaluated the neutralizing potency of the 43 generated mAbs. In *TNFRSF13B* mutation carriers, increased VH mutation frequency correlated with lower IC_50_ values, indicative of greater neutralization potency. This relationship was not observed in mAbs derived from WT subjects (Figure 4D).

Given that highly mutated mAbs isolated from subjects with TNFRSF13B variants neutralized Wuhan-Hu1 spike-pseudotyped viruses more effectively than antibodies obtained from subjects with the WT *TNFRSF13B* genotype, we hypothesize that the interactions between these antibodies and their targets may help explain the observed neutralization efficacy.

### Antibodies derived from subjects with TNFRSF13B variants have more points of contact with the RBD of the Wuhan-Hu-1 spike protein than antibodies derived from subjects expressing WT TNFRSF13B

We observed that plasma from subjects with TNFRSF13B variants exhibited increased neutralization of pseudotyped Wuhan-Hu-1 virus. We next asked if these functional differences could be seen due to enhanced binding to the receptor-binding domain (RBD) of the spike protein. To further investigate, we utilized light and heavy chain sequences from monoclonal antibodies sequenced from 6 subjects expressing WT *TNFRSF13B* and from 5 subjects carrying mutant alleles (1 homozygous P251L, 2 heterozygous P251L, 1 heterozygous C104R, and 1 heterozygous G190R).

Using ABodyBuilder (Kenlay et al., 2024), we produced a total of 55 and 46 monoclonal antibody structures derived either from subjects with WT *TNFRSF13B* or mutant genotypes. We then docked those antibodies to Wuhan-Hu-1 spike protein (PDB: 6VYB (Walls et al., 2020)) using ClusPro 2.0 (Desta et al., 2023). The resulting complexes were then analyzed with PRODIGY (PROtein binDIng enerGY prediction) (Stranges and Kuhlman, 2013) to estimate binding affinity (ΔG) based on interfacial residue contacts and surface properties. Using thermodynamic relationships, ΔG was also converted into a dissociation constant (K_d_). PRODIGY results showed comparable binding affinity to Wuhan-Hu-1 spike protein and comparable dissociation constants of antibodies obtained from subjects with WT *TNFRSF13B* or from subjects with *TNFRSF13B* mutant genotypes (Extended Data Fig. 3A-B).

Despite comparable binding affinities, structural mapping through PyMOL (PyMOL) showed that antibodies obtained from subjects with *TNFRSF13B* mutant genotypes, particularly those carrying the P251L variant, bound predominantly to the RBD of the spike protein and most of N-terminal domain (NTD), whereas antibodies from subjects with WT *TNFRSF13B* exhibited a more dispersed binding footprint that included the subunit S2 regions (Figure 5A). The top 60 residues with the largest absolute differences in antibody-spike contact frequency between antibodies from WT and TNFRSF13B variant subjects are listed in Extended Data Tables 3, 4, and 5. Furthermore, representative structures of antibody-RBD complexes with the highest number of residue contacts demonstrate that antibodies obtained from subjects with *TNFRSF13B* mutations engage more RBD residues than antibodies obtained from subjects with WT *TNFRSF13B* (Figure 5D). In addition to evaluating the antibody contact frequencies across the spike protein, we analyzed contact distances for residues contacted by both WT and mutant antibodies. At these shared sites, antibodies from *TNFRSF13B* mutant subjects, particularly P251L and G190R carriers, exhibited shorter contact distances to RBD residues (Figure 5C).

**Figure 5:**
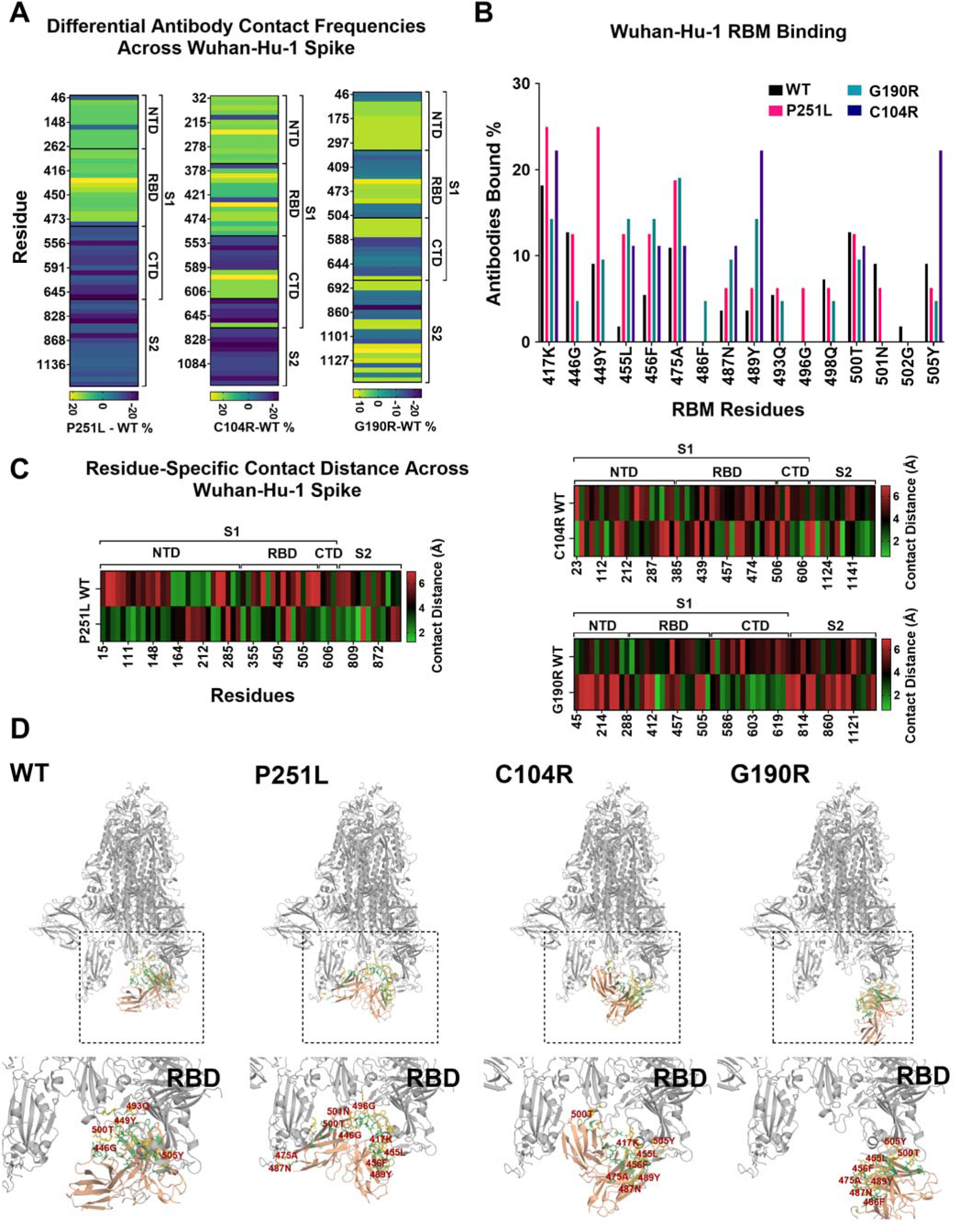
*In silico* comparison of WT and *TNFRSF13B* mutant-derived antibodies binding to the SARS-CoV-2 Wuhan-Hu-1 spike protein. Heavy and light chain sequences from WT and *TNFRSF13B* mutant subjects were modeled with ABodyBuilder, docked to the Wuhan-Hu-1 spike protein (PDB: 6VYB) using ClusPro. In total, 55 monoclonal antibodies from 6 WT subjects and 46 from 5 mutant subjects (MUT) were analyzed. **A.** Heatmap showing the top 60 SARS-CoV-2 spike residues with the largest absolute difference (|Δ MUT-WT|) in contact frequency between MUT and WT antibodies. Contacts were defined as atoms within 4 Å between antibody and spike residues. The color scale indicates the difference (MUT-WT %) in contact frequency where yellow/green indicates higher contact frequency in MUT antibodies and blue indicates higher frequency in WT antibodies. Residues are grouped by spike domain (NTD; N-terminal domain, RBD; receptor-binding domain, CTD; C-terminal Domain, subunit S2), with every third residue labeled to improve clarity. **B.** Comparison of WT and MUT antibodies binding to residues within the receptor-binding motif (RBM) of the SARS-CoV-2 spike proteins. Bars present the percentage of antibodies contacting each residue within 4 Å. **C.** Heatmaps show residue-level median antibody–spike binding distances at shared Wuhan-Hu-1 spike residues for WT and mutant antibodies. The color scale indicates contact distance (Å), with green representing tighter (closer) contact and red representing looser (more distant) contact. **D.** Representative images of WT and MUT antibody complexes with highest number of contacts with SARS-CoV-2 RBD visualized in PyMOL. RBD is shown as a gray ribbon, and the antibody variable domain is shown as an orange ribbon. Residues forming contacts within 4 Å are displayed in a ball-and-stick representation, with green indicating paratope residues (on the antibody) and yellow indicating epitope residues (on the RBD). Key interacting residues are labeled red.

Structural studies of RBD-ACE2 (Angiotensin-Converting Enzyme 2) complexes have established that residues Leu455, Phe456, Phe486, Asn487, Tyr489, Gln493, Gln498, Asn501, and Tyr505 sit within the receptor-binding motif (RBM) and directly contact ACE2, making them important neutralization epitopes (Greaney et al., 2021; Lan et al., 2020; Starr et al., 2020). When we examined these positions specifically, antibodies of subjects with *TNFRSF13B* mutations bound these RBM residues, along with other additional RBM residues, more frequently than WT antibodies as well (Figure 5B).

To further assess whether antibody neutralization potency was associated with preferential engagement of specific regions of the spike protein, antibodies were stratified into higher potency (IC_50_ ≤ 0.01 µg/mL) and lower potency (IC ≥ 0.01 µg/mL) groups. Despite being classified as lower potency for this specific analysis (IC_50_ ≥ 0.01 µg/mL), the majority of antibodies with IC_50_ ≥ 0.01 µg/mL fall within IC_50_ ranges reported as effective neutralization in other SARS-CoV-2 antibody studies (Chen et al., 2025; Liu et al., 2020).

We found that 4 of 19 WT monoclonal antibodies (21.1%) and only 2 of 20 (10%) monoclonal antibodies from subjects with TNFRSF13B variant are in the lower potency groups. We also found that higher potency antibodies had tighter average contact distance to RBD residues (6.6 Å) compared to lower potency antibodies (7.4 Å) (Extended Data Fig. 3C). Thus, the *in silico* analyses suggest that antibodies obtained from subjects with TNFRSF13B variants are biased toward RBD of Wuhan-Hu-1 spike protein, which is consistent with more favorable virus neutralization.

### Subjects with TNFRSF13B variants produce IgG with decreased sialic acid, terminal galactose, and core fucose

IgG post-translational modifications, particularly Fc glycosylation, determine inflammatory functions of antibodies. Decreased IgG galactosylation and sialyation were found in prevalent inflammatory and autoimmune diseases like rheumatoid arthritis (RA) (Gindzienska-Sieskiewicz et al., 2016; Ohmi et al., 2016), systemic lupus erythematosus (SLE) (Vuckovic et al., 2015), inflammatory bowel disease (IBD) (Gaifem et al., 2024; Simurina et al., 2018), and multiple sclerosis (MS) (Wuhrer et al., 2015). IgG Fc glycosylation is a critical modulator of antibody effector functions; those functions include complement activation and Fc receptor engagement (Borrok et al., 2012; Raju, 2008; Spiteri et al., 2021). Furthermore, IgG antibodies that do not contain core fucose at Asn 297 glycan have significantly higher affinity to immune effector cells such as natural killer (NK) cells and macrophages (Golay et al., 2022; Okazaki et al., 2004; Shields et al., 2002).

Ash et. al (Ash et al., 2022) reported that glycan profile of bulk total IgG predicts COVID-19 severity; specifically, decreased IgG sialyation and galactosylation are more often observed in hospitalized subjects compared to non-hospitalized subjects. We hypothesized that the increased propensity for severe disease in subjects with *TNFRSF13B* mutant genotypes might be owed to differences in antibody effector functions brought about by IgG Fc glycosylation.

To evaluate if changes in IgG-Fc glycan structures contribute to disease severity in individuals carrying *TNFRSF13B* missense mutations, we isolated total IgG from plasma of subjects with WT *TNFRSF13B* or P251L *TNFRSF13B*. Using lectin-based assays, we quantified specific glycan residues on purified and equivalent amounts of IgG immobilized on ELISA plates. Biotinylated Ricinus communis agglutinin I (RCA-1) was used to detect terminal β-linked galactose residues, Sambucus nigra agglutinin (SNA-EBL) identified α2,6-linked sialic acid residues, and Lens Culinaris Agglutinin (LCA) was used to detect α1-6 fucose attached to GlcNAc near asparagine.

We observed that subjects with the P251L TNFRSF13B (n=22) and severe SARS-CoV-2 infection had less IgG sialic acid and terminal galactose compared to those with the WT *TNFRSF13B* (n=40) (Figure 6A-B). Furthermore, the plasma of subjects with P251L *TNFRSF13B* (n=18) also had slightly less IgG core fucose in relation to IgG in plasma of subjects with the WT *TNFRSF13B* (n=19) (Figure 6C).

**Figure 6:**
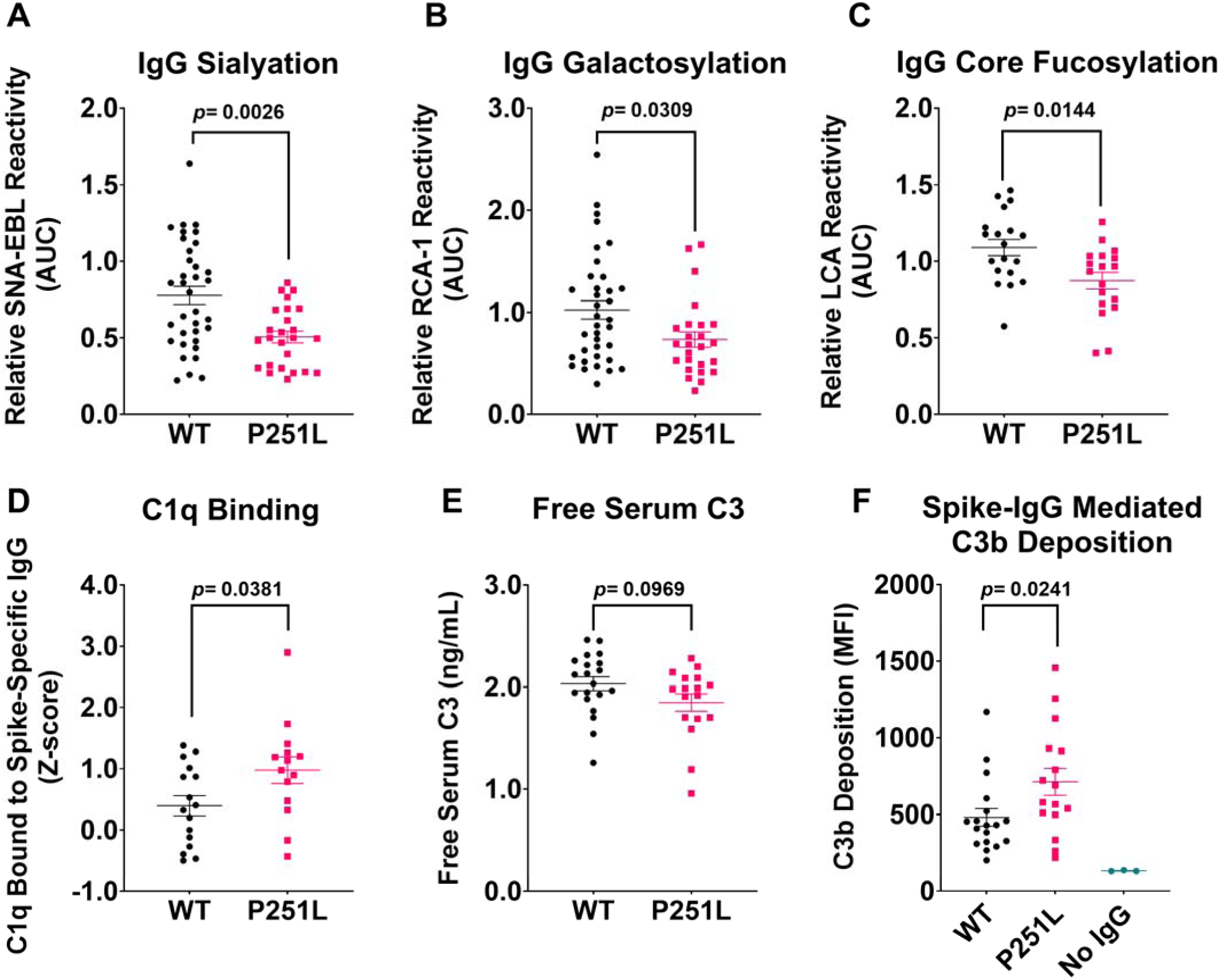
*TNFRSF13B* genotype alters IgG glycosylation and complement activation. **A-C** IgG sialic acid, terminal galactose, and core fucose, were measured in subjects with WT *TNFRSF13B* (A and B: n= 40, C: n= 19) and subjects with P251L *TNFRSF13B* (A and B: n= 22, C: n= 18). Concentrations of input purified plasma IgG were normalized to 25 µg. Lectin reactivity was quantified by obtaining OD405nm absorbance values from decreasing serial dilution in ELISA. The absorbance values were then calculated as area under the curve (AUC) determined in GraphPad Prism. To mitigate for inter-plate variation, AUC values were normalized to an internal reference sample included on each plate. **D.** Spike-specific IgG-C1q interaction was assessed by ELISA with 0.5 mg of purified IgG bound to spike protein before incubating with human C1q protein (WT, n= 16; P251L, n= 14). OD405nm absorbance values were obtained, and z-scores were calculated to account inter-plate variation. **E.** Plasma C3 concentrations were measured in WT and P251L carriers using Abcam C3 ELISA kit (WT, n= 20; P251L, n= 17). **F.** Spike-specific IgG mediated C3b deposition was determined by incubating spike-RBD-coated beads with 0.5 µg of spike RBD-specific IgG (determined separately by ELISA) from subjects with WT (n=18) or TNFRSF13B P251L variant (n=16), followed by antibody-depleted serum as a complement source. C3b deposition was measured by flow cytometry and shown as mean fluorescence intensity (MFI). Comparisons between groups were done using Mann-Whitney test for **(A-C and F)** unpaired t-test for **(D-E)** in GraphPad Prism (v10).

Multivariable linear regression analyses adjusting for demographics and clinical covariates indicated that differences in IgG glycosylation were associated with the *TNFRSF13B* genotype. After accounting for age, sex, body mass index (BMI), comorbidity status, and time of sample collection after COVID-19 diagnosis, the association between TNFRSF13B variants and reduced IgG sialylation and galactosylation remained, with β coefficients of −2.84 and −4.53, respectively (IgG sialylation; *p*= 0.051, IgG galactosylation; *p=* 0.001, Extended Data Table 6).

While IgG Fc galactosylation is required for IgG hexamerization and C1q recruitment (Peschke et al., 2017; van Osch et al., 2021), IgG Fc galactosylation also blocks complement mediated-inflammation (Karsten et al., 2012). Moreover, sialylation of IgG Fc domain reduce binding to C1q, deposition of C3b and activation of complement-mediated cytotoxicity (Quast et al., 2015). Because subjects with P251L TNFRSF13B variants had less sialic acid and galactose on IgG antibodies, our results indicate that the *TNFRSF13B* genotype determines the IgG glycan profile and suggests a mechanism for enhanced complement mediated inflammation and severe SARS-CoV-2 infection.

### Heightened complement activation in subjects with severe SARS-CoV-2 infection carrying the P251L TNFRSF13B variants

Heightened complement activation may contribute to the severe outcomes following SARS-CoV-2 infection in subjects with *TNFRSF13B* mutant genotypes given the increased IgG1 and IgG3 isotypes and reduced IgG Fc glycosylation. As a surrogate for IgG-dependent complement activation, we first investigated complement factor C1q binding to spike-specific IgG. C1q binding to IgG immobilized on SARS-CoV-2 spike RBD protein was detected using an anti-human C1q antibody by ELISA. This strategy ensured the proper spatial orientation and Fc conformation of IgG, and more accurately reflects physiologic interactions of spike-specific antibodies with antigen. We observed increased C1q recruitment by spike-specific IgG from P251L carriers compared to spike-specific IgG from subjects with WT TNFRSF13B, (Figure 6D), indicating that *TNFRSF13B* mutant genotypes enhance IgG-dependent C1q complement activation.

Consistent with the observation of increased complement activation, we also quantified circulating serum C3 levels and observed a modest reduction in P251L carriers compared to subjects with WT TNFRSF13B (1.84 µg/mL vs. 2.03 µg/mL, median), suggesting the possibility of increased complement consumption in subjects with the P251L variant (Figure 6E). To directly measure IgG-mediated C3 activation, equal amount of spike-RBD specific IgG was incubated with spike-RBD coated beads, followed by incubation with antibody depleted human serum as a complement source. C3b deposition on spike RBD beads was then measured by anti-C3b antibody and flow cytometry.

Spike-RBD specific IgG from subjects with P251L variant had increased C3b deposition on the spike-RBD beads compared to IgG from WT subjects (Figure 6F). Importantly, hardly any C3b signal was detected in the absence of IgG, indicating that C3b deposition measured in this assay was IgG dependent and predominantly through the classical pathway which is initiated by C1q binding to the Fc regions of antigen-bound IgG.

Because multiple antibody properties were evaluated, *p* values were adjusted for multiple comparisons using the Benjamini–Hochberg false discovery rate method. The adjusted *p* values (q-values) are: IgG sialylation q = 0.013, IgG galactosylation q = 0.039, IgG fucosylation q = 0.036, IgG-mediated C1q binding q = 0.038, and IgG-mediated C3b deposition q = 0.024. All primary findings remained significant after correction. Therefore, the data suggests that the *TNFRSF13B* mutant genotypes enhance disease severity by augmenting antibody-dependent complement activation in response to infection.

## Discussion

Genetic studies of subjects with COVID-19 have uncovered association between severe disease and variants in genes involved in viral entry, including ACE2-TMPRSS2 cell surface receptor complex (Ren et al., 2022; Vadgama et al., 2022), cytokine production (Ishak et al., 2022; Strafella et al., 2020), and genes within the human leukocyte antigens gene complex (HLA) (Amoroso et al., 2021; Pisanti et al., 2020; Sakuraba et al., 2020). In contrast, the impact of genes controlling antibody responses in severe COVID-19 and ARDS is not well known. We reasoned that the COVID-19 pandemic offered a unique opportunity to appreciate the contributions of common variants in *TNFRSF13B*, a highly polymorphic gene involved in B cell differentiation and antibody production. Schmidt et. al (Schmidt et al., 2021) reported that a child with lethal COVID-19 carried both a homozygous deletion in TANK-binding kinase 1 (TBK1) gene and homozygous C104R TNFRSF13B variant, suggesting that TNFRSF13B variants that impair the function of the receptor may contribute to the severity of SARS-CoV-2. Consistent with the idea that TNFRSF13B variants are associated with severity of disease following SARS-CoV-2 infection, we found that certain TNFRSF13B variants including the P251L allele are associated with severe SARS-CoV-2 infection and ARDS.

The TNFRSF13B variant encoding leucine instead of a proline in the intracellular domain of the receptor (the P251L variant) is not thought to impair receptor signaling (Salzer et al., 2005) or to be associated with immunodeficiency (Karaca et al., 2018). However, we found the P251L variant to be a risk factor for ARDS following SARS-CoV-2 infection. *In silico* predictions yield contradicting results as PolyPhen-2 (Adzhubei et al., 2010) predicts P251L allele to be “possibly damaging” (Score of 0.728) of the protein structure and function. In contrast, ClinVar finds no links of P251L to phenotypic traits or disease and classifies P251L as benign (Landrum et al., 2018). The P251L variant is found in 11.6% of individuals or more depending on ethnicity. While 38% of East Asians have the P251L allele, only 10% of Europeans express the P251L allele, per GnomAD v4.1 (Chen et al., 2024). Importantly, the high frequency of the P251L allele in the population is not consistent with a detrimental impact on health.

In accord with variant-induced protective functions, subjects with the P251L variant exhibited modestly enhanced neutralization of the Wuhan-Hu-1 virus. Although the sample size was smaller, we also observed stronger neutralization against the Delta strain, suggesting a broader serum neutralization capacity. Furthermore, based on *in silico* analyses, antibodies obtained from subjects expressing TNFRSF13B variants bound to more residues within the receptor-binding motif of the Wuhan-Hu-1 spike proteins than antibodies obtained from subjects with WT TNFRSF13B. These findings suggest that common TNFRSF13B variants might augment antibody-mediated host defense against mutable viruses.

Severe SARS-CoV-2 infection is thought to be driven by host inflammatory responses rather than by uncontrolled viral dissemination (Del Valle et al., 2020; Hu et al., 2021; Lucas et al., 2020). The pathophysiology of ARDS highlights the lung’s vulnerability to excessive inflammation and complement activation causing irreversible damage to lung alveolar epithelial and endothelial cells (Detsika et al., 2024; Gralinski et al., 2018; Ma et al., 2025; Zhou et al., 2025). Our results suggest that the increased propensity for ARDS in subjects with TNFRSF13B variants is linked to reduced IgG Fc glycosylation and enhanced complement activation, consistent with prior studies demonstrating that IgG Fc glycan composition regulates complement function (Frischauf et al., 2024) (Dekkers et al., 2017; Diebolder et al., 2014; Karsten et al., 2012; Kristic and Lauc, 2024; Quast et al., 2015). Additionally, multivariate genome-wide association studies (Sharapov et al., 2025; Shen et al., 2017) examining IgG *N*-glycosylation loci found TNFRSF13B P251L variant (rs34562254) to be associated with decreased IgG sialylation and galactosylation, supporting our findings that *TNFRSF13B* controls IgG glycosylation. Given that SARS-CoV-2 targets the lungs, antigen-specific antibodies may direct complement activation to the lung increasing severity of manifestations and likelihood of progression to ARDS. Consistent with this mechanism, we found previously that TNFRSF13B variants including P251L increase the risk for antibody-mediated rejection in kidney transplant recipients by 5-fold, which is often due to complement-mediated injury (de Mattos Barbosa et al., 2021a).

While our findings provide new insights into how TNFRSF13B variants determine outcomes of SARS-CoV-2 infection, several limitations should be noted. This study was limited by the relatively small cohort size. In addition, although lectin-based ELISA allowed quantification of overall IgG glycan abundance, we were unable to differentiate between Fc and Fab associated glycosylation, despite Fab glycosylation being a smaller fraction of total IgG glycans (van de Bovenkamp et al., 2016).

In conclusion, our data show that *TNFRSF13B* genotypes dictate IgG glycosylation in response to infection, which in turn regulates complement activation and balances pathogen clearance with tissue repair. Although certain variants are linked to ARDS, they also enhance viral neutralization and possibly reduce transmission, underscoring an evolutionary tradeoff that likely preserves these *TNFRSF13B* alleles in human populations.

## Materials and Methods

### Human Participants

Research was approved by the University of Michigan Medical School, Henry Ford Health System and University of Louisville Institutional Review Boards. Written informed consent was obtained from all study participants prior to collection of any samples and inclusion to the study. A total of 108 patients with polymerase chain reaction positive for SARS-CoV-2 were consented after COVID-19 diagnosis and treated as outpatients or hospitalized in the acute phase of disease. Patients were recruited at the University of Michigan Hospital (n= 22) and at the Henry Ford Health System (n= 7). Additionally, DNA, whole blood, and plasma samples were obtained from the University of Michigan (n= 58) and University of Louisville (n= 21) sample biorepositories. Most samples (94/108) were collected between April and September 2020 when mostly the wild-type strain was in circulation. No virus genotyping data was available for the patients recruited for the study.

Patients were of both genders of 18 years of age and older, individuals coinfected with HIV, Hepatitis B or C were excluded. Median age was 50 years (range 24-88), 42% were females and 58% were males. Many of the subjects had comorbidities such as diabetes, obesity, cardiovascular disease, hypertension, or cancer. When possible, for data analysis, patients characteristics, age, sex, BMI, sample collection time from COVID-19 diagnosis, and comorbidities are accounted by various statistical tests. The subjects were classified as severe based on criteria that include symptoms such as pneumonia, acute respiratory distress syndrome (ARDS), acute hypoxemic respiratory failure (AHRF), UCI admission, and necessity for intubation or extracorporeal membrane oxygenation (ECMO).

Plasma was separated from heparinized blood. For neutralization assays, the plasma samples were heat-inactivated at 56°C for 1 hour prior to the start of the assay. Peripheral blood mononuclear cells (PBMCs) were isolated with Ficoll-Paque PLUS density gradient media (Cytiva, Cat#17-1440-02). Genomic DNA was extracted from PBMCs or whole blood using DNeasy Blood & Tissue Kit following the manufacturer directions (Qiagen Cat#69504). Plasma and DNA were frozen and stored in a -80°C freezer while PBMCs were stored in the liquid nitrogen until posterior use.

### *TNFRSF13B* sequencing and analysis

Located on chromosome 17, *TNFRSF13B* is composed of 5 exons spanning 34Kb. We amplified *TNFRSF13B* exons 1, 2, 3, and 4 with primers previously described by Salzer and colleagues *(11)*. Exon 5 was amplified with a pair of primers previously described by us *(17)*. Polymerase chain reaction was performed with Taq DNA Polymerase, native (Thermo Fisher Scientific Cat#18038-042) at 95°C for 15 minutes, 46 cycles at 94°C for 30 seconds, 60°C for 15 seconds, 72°C for 30 seconds, followed by 72°C for 5 minutes. DNA bands were visualized in 2% agarose gels and 10 µL of the PCR products were purified with ExoSAP-IT PCR Product Cleanup Reagent (Applied Biosystems Cat# 78202) according to the manufacturer instructions before sequencing. Sanger sequencing was performed at Eurofins Genomics facilities and sequences were aligned using Sequencher 5.4.6 software (Gene Codes Corporation, Ann Arbor, MI).

### Enzyme-Linked Immunosorbent Assay for Detection of Human Immunoglobulins

Nunc MaxiSorp ELISA plates (Thermo Fisher Scientific Cat# 44-2404-21) were coated overnight with goat anti-human IgM (SouthernBiotech Cat#2020-01, RRID:AB_2795599), IgA (SouthernBiotech Cat#2050-01, RRID:AB_2795701) or IgG (SouthernBiotech Cat#2040-01, RRID:AB_2795640) diluted at 4 µg/mL in PBS. Plates were blocked and incubated with plasma from COVID-19 convalescent subjects. Bound Igs were detected by adding 4 µg/mL HRP-conjugated goat anti-human IgM (SouthernBiotech Cat#2020-05, RRID:AB_2795603), IgA (SouthernBiotech Cat#2050-05, RRID:AB_2687526) or IgG (SouthernBiotech Cat#2040-05, RRID:AB_2795644) diluted in PBS. Reactions were visualized by subsequent addition of 2,2′-Azino-bis (3-ethylbenzthiazoline-6-sulfonic acid) substrate (SouthernBiotech Cat#0202-01). Readings were recorded at 405 nm with an Agilent Biotek Plate Reader.

### Spike-specific B cell panning and single cell V(D)J analysis

SARS-CoV-2 spike protein S1 subunit-specific B cells were isolated from COVID-19 convalescent subjects PBMCs as previously described *(17)*. Briefly, PBMCs were thawed and cultured for 72 hours at 37°C, 5% CO_2_ in RPMI 1640 (Gibco, Cat#11875093) supplemented with L-Glutamine, 10% FBS, 100 U/mL Pen Strep, 0.1% 2-mercapthoethanol, 5μM CpG ODN 2006, 10 ng/mL CD40L, 50 ng/mL IL-2, 2 ng/mL IL-15, and 10 ng/mL IL-21 (cytokine cocktail media). Cells were retrieved after 72 hours incubation, resuspended in fresh cytokine cocktail media and transferred to wells coated with rabbit anti-Avi-tag antibody (4 µg/mL, GenScript, Cat#A00674, RRID: AB_915553) bound to S1-C-6HIS-Avi (4 µg/mL, ABclonal, Cat#RP01261) previously blocked with PBS 10% FBS for 24 hours. Wells were washed to remove unbound cells by 3 washes, the media was replaced and then cells were incubated at 37°C 5% CO_2_ for additional 48 hours. Bound cells were collected by vigorous pipetting.

Up to 10,000 cells were analyzed by Chromium Next Gel Bead-in-Emulsions (GEM) Single Cell V(D)J Technology. Briefly, cells were identified via generation of GEMs by combining barcoded Single Cell V(D)J 5’ Gel Beads v1.1, a master mix with cells (Chromium Next GEM Single Cell 5′ Library and Gel Bead Kit v1.1; 10x Genomics, Cat#1000165), and partitioning oil on Chromium Next GEM Chip G (Chromium Next GEM Chip G Single Cell Kit; 10x Genomics, Cat#1000120) and reverse transcription and cDNA amplification were performed as recommended by the manufacturer. Next, the targeted enrichment from cDNA was conducted with the Chromium Single Cell V(D)J Enrichment Kit, Human B Cell (10x Genomics, Cat#1000016). The cDNA quality control (QC) analysis was carried out in an Agilent 2100 Bioanalyzer (Agilent Technologies, Santa Clara, CA) using the Agilent High Sensitivity DNA Kit (Agilent Technologies, Cat#5067-4626). The V(D)J enriched library was then constructed via Chromium Single Cell 5’ Library Construction Kit v1.1 (10x Genomics, Cat#1000166) and libraries were sequenced in a NovaSeq™ 6000 Sequencing System (Illumina, San Diego, CA). Chromium Single Cell RNA-seq output was processed in the Cell Ranger pipelines (10x Genomics, Pleasanton, CA, RRID:SCR_017344) and the V usage and clonotype profiles were generated and visualized by Loupe VDJ Browser. Sequences were paired and quality controlled with pRESTO (, as described in *(24, 25)* and annotated with IgBLAST *(26)* using default parameters and The International Immunogenetics Information System (IMGT) reference database (RRID:SCR_011812) *(24, 25, 27, 80 ,83)*. The resulting data were imported into ImmuneDB *(28)* for further downstream analyses.

### Quantification of spike-specific IgG antibodies

MaxiSorp plates were coated with 0.2 µg/well of SARS-CoV-2 Spike Protein S1/S2 (aa11-1208) (Thermofisher, catalog #RP-87680) or SARS-CoV-2 Spike Protein RBD (Thermofisher, catalog # RP-87704) overnight at 4°C. Following coating, plates were blocked with 2% BSA solution for one hour at room temperature. Subjects’ plasmas were diluted at 1:100 and 100 µL of diluted plasma was used. After one hour of incubation at room temperature, bound IgG to the coated spike protein was detected using 4 µg/mL goat anti-human IgG HRP (SouthernBiotech Cat#2040-01, RRID:AB2795640). Reactions were visualized by subsequent addition of 2,2′-Azino-bis (3-ethylbenzthiazoline-6-sulfonic acid) substrate (SouthernBiotech Cat#0202-01). Readings were recorded at 405 nm with an Agilent Biotek Plate Reader.

### Neutralization assays

Neutralization assays were performed as previously described *(17)*. Briefly, 293T-ACE2 cells were cultured into 96-well plates to obtain 50% confluence on the day of transduction. Heat-inactivated COVID-19 convalescent subjects plasma were serially diluted into DMEM (Gibco Cat#11965092) 10% FBS (Gibco Cat#10082-147) 1X Glutamax (Gibco Cat#35050-061) 100 U/mL Pen Strep (Gibco Cat#15140-122) and incubated with lentivirus pseudotyped with Wuhan-Hu-1 SARS-CoV-2Δ19AA spike (2.66 x 10^5^ TU/mL) and Delta variant spike (B.1.617.2; 3 x 10^45^ TU/mL; BPS Bioscience Cat#78215) for 30 minutes at room temperature. Cells were transduced with the virus-plasma solution or virus alone and incubated at 37°C 5% CO_2_ for 72 hours. Luminescence was detected 72 hours post-transduction by luciferase assay (Bright-Glo Luciferase Assay System, Promega Cat#E2620) according to the manufacturer instructions. Luminescence was detected in 96-well white plates by Synergy 2 plate reader (Biotek Instruments, Winooski, VT). Neutralization curves were generated and ID_50_ and ID_80_ values were obtained using GraphPad Prism analysis feature: “[Inhibitor] vs. response – Variable slope (four parameters)”.

### In silico analysis

After isolating the monoclonal antibodies from subjects with SARS-CoV-2 infection, the monoclonal antibodies were sequenced to identify the light and heavy chain sequences using the protocol described above. Using ABodyBuilder *(31)*, the sequences of the light and heavy chains were imported to create a total of 55 WT and 46 *TNFRSF13B* variant monoclonal antibody structures. For the resulting antibody structures, glycans, metal ions, and solvent molecules were removed using PYMOL. The antibody structures were then docked to Wuhan-Hu-1 spike protein (PDB: 6VYB; RCSB.org). PDB 6VYB is a prefusion Wuhan-Hu-1 SARS-CoV-2 spike cryo-EM structure. Using ClusPro 2.0 *(33)* and the antibody docking mode feature, the antibody structures are docked to Wuhan-Hu-1 spike protein. The resulting antibody-spike complexes were then analyzed with PRODIGY *(34)* to estimate binding affinity (ΔG) and dissociation constant (K_d_).

Further structural analyses were performed using PyMOL*(35)* with custom Python scripts through the PYMOL console. For each antibody-spike complex, spike residues within 4 Å of any antibody atom were identified and recorded as antibody-spike contacts. This dataset was then used to calculate, for each spike residue including those in the receptor binding motif, the proportion of antibodies from WT and *TNFRSF13B* variant groups that contacted the residue. Differences in the binding frequency between MUT and WT antibodies were calculated for each spike residue, and the 60 residues showing the largest absolute differences were selected for visualization as a heatmap. These residues were also used to generate the contact distance (up to 7 Å) heatmap using a custom PYMOL script.

### Quantification of IgG sialyation, galactosylation, and core fucosylation

IgG was purified from human serum using the Melon Gel IgG Spin Purification Kit (Thermo Fisher, Cat# 45212). Following purification, IgG purity was confirmed by performing ELISA and SDS-PAGE to check for the presence of IgM and IgA. The purified IgG was quantified by ELISA, and 25 µg of IgG was used to coat each well in a serial dilution manner of a MaxiSorp 96-well plate and the plates were incubated overnight at 4°C. The following day, the plates were washed three times with PBS and then blocked with by adding 200 µL of Carbo-Free Blocking Solution (Vector Laboratories. SKU# SP-5040-125) per well for 1 hour at room temperature. After blocking, the samples were incubated with a divalent ion solution (1 mM CaCl_2_, 1 mM MgCl_2_, 1 mM MnCl_2_) for 15 minutes at room temperature to optimize lectin-IgG binding. Biotinylated lectins RCA-1, SNA-EBL, LCA (Vector Laboratories, SKU# B-1085-1, B-1305-2, B-1045-5) were diluted to 5 µg/mL in Carbo-Free Blocking Solution, and 100 µL of the diluted lectins was added to each well, followed by incubation for 1 hour at room temperature. Signal detection was carried out by adding 6.25ng of Streptavidin-HRP (Thermo Fisher, Cat# N100) to each well and incubated at room temperature for 45 minutes. For color development, 50µL of 1-step ABTS solution (Thermo Fisher, Cat# 37615) was added to each well, and absorbance was measured at 405nm with a Biotek luminescence plate reader.

Lectin binding was determined by ELISA across serial dilutions. Area under the curve (AUC) was calculated using the trapezoidal method from background subtracted absorbance values using GraphPad Prism and normalized to a same reference internal sample (Purified Human IgG, SouthernBiotech, Cat. No: 0150-01) included on each plate to correct for inter-plate variability.

### Measuring Complement C3 in Human Serum

Following the manufacturer’s instructions for Abcam Human Complement C3 ELISA kit (Abcam, ab108823), patient plasma was diluted 1:125,000. Briefly, 50µL of complement C3 standard or diluted plasma samples were added to the microplate strips provided in the kit. 200µL of the antibody cocktail was added to the samples and incubated for two hours at room temperature. Plates were developed with developer solution and read at 450nm using an Agilent Biotek Plate Reader.

### Quantification of spike-specific IgG-complement C1q interaction

Maxisorp plates were coated with 0.2 µg/well of SARS-CoV-2 Spike Protein S1/S2 (aa11-1208) (Thermofisher, catalog #RP-87680) overnight at 4°C. Following coating, plates were blocked with 2% BSA solution for one hour. Then, serial dilution starting from 0.5mg of purified IgG from patient samples was added per well and incubated at room temperature for one hour. Following incubation with spike protein and purified IgG, 1 µg of recombinant purified human C1q protein (Complement Technology, Catalog # A099) in PBS with 1mM of CaCl_2_, and 1mM of MgCl_2_ was added to each well and incubated for 1 hour at room temperature. After 1 hour incubation with human C1q, plates were washed 3 times with PBS and anti-human C1q biotinylated (ThermoFisher, MA1-40312) was added to each well in the dilution of 1:50. After one hour incubation at room temperature, 6.25 ng of HRP-conjugated Streptavidin-HRP (ThermoFisher, N100) was added per well for color development using 50µL of 1-step ABTS solution (Thermo Fisher, Cat# 37615). Absorbance values were measured at 405 nm using a Biotek Luminescence Plate Reader.

To account for inter-plate variability, absorbance values from all ELISA plates are combined into a single matrix and global mean and standard deviation of the full dataset is calculated. For each absorbance value “x”, z-score was calculated using the formula, Z= (x – global mean)/standard deviation.

### Measuring C3b deposition

SARS-CoV-2 spike RBD-coupled magnetic beads (AcroBiosystems, MBS-K002) were reconstituted to final concentration of 1mg beads/mL. For each reaction, 10µL of spike RBD beads were incubated with 0.5µg of spike RBD-specific IgG for 45 minutes at room temperature. The concentrations of spike RBD-specific IgG in patients plasma samples were determined by ELISA separately as described above. Following incubation, IgG bound spike-RBD beads were washed 3 times with PBS. As a complement source, 30 µL of antibody depleted human serum (Pel-Freez, 34041-1) in 1mL of GVB++ buffer with Ca^2+^ and Mg^+2^ (Complement Technology, Catalog# B102) was added to the IgG bound spike-RBD beads and incubated for 30 minutes at 37°C. After complement incubation, beads were washed 3 times with PBS and stained with 1µL of anti-C3/C3b/iC3b PE (BD Pharmingen, Catalog #567667). Following staining, beads were analyzed by flow cytometry using a BD LSR flow cytometer.

## Statistics

All comparisons were completed with GraphPad Prism10 software (GraphPad Software version 10.5.0, La Jolla, CA, RRID:SCR_002798). Detailed statistics for each panel can be found on the table and figure legends. To determine if any observed missense mutations were associated with SARS-CoV-2 infection and/or COVID-19 severity we used Fisher’s exact test to compare the number of mutated and wild type alleles in COVID-19 subjects versus the numbers reported for the general population according to the GnomADv4.1(Chen et al., 2024). Relative risk was calculated by the Koopman asymptotic score. Averages were compared by unpaired t test or nonparametric Mann-Whitney test. *p* value of equal or less than 0.05 was considered significant.

## Supplemental Material

Includes data showing serum immunoglobulin concentrations in subjects in accord to disease severity and ARDS; graphing of predicted K_d_ and delta G of spike-specific monoclonal antibodies according to genotype of the subject they were isolated from; data depicting the clinical and demographic characteristics of patients in the severe SARS-CoV-2 infection group; data depicting the distribution of the *TNFRSF13B* missense mutation P251L across ethnic groups in subjects with SARS-CoV-2 infection; data depicting the distribution of top 60 residues of anti-spike antibodies obtained from WT or P251L subjects that differ the most in contact frequency with the RBD; data depicting the distribution of top 60 residues of anti-spike antibodies obtained from WT or C104R subjects that differ the most in contact frequency with the RBD; data depicting the distribution of top 60 residues of anti-spike antibodies obtained from WT or G190R subjects that differ the most in contact frequency with the RBD. We also show multivariable linear regression analysis of IgG sialylation and galactosylation.

Extended Data Figs. 1-3

Extended Data Tables 1-6

## Data availability statement

All data are available in the main text or in the supplementary materials. Data on human subjects that is not included in the manuscript will be made available de-identified upon reasonable request.

## Data Availability

All data produced in the present work are contained in the manuscript

## Acknowledgments

We acknowledge the University of Michigan Medical School Central Biorepository, the Department of Medicine and the Division of Infectious Diseases; the University of Louisville Respiratory Biorepository Program and Dr. Christina L Kaufman, Associate Professor in the Department of Cardiovascular and Thoracic Surgery at the University of Louisville for the samples provided. We acknowledge the Richard J. Fasenmyer Professorship in Immunopathogenesis at Case Western Reserve School of Medicine.

## Abbreviations

ARDS: Acute Respiratory Distress Syndrome
Ig: Immunoglobulin
TNFRSF13B: Tumor Necrosis Factor Receptor Super Family 13B
ICU: Intensive Care Unit
SARS-CoV-2: Severe Acute Respiratory Syndrome Coronavirus 2
RBD: Receptor Binding Domain
ACE: Angiotensin-Converting Enzyme

## Author contributions

Conceptualization: LN, MGMB, JLP, MC

Methodology: LN, MGMB, IC, MB, JPL, MC

Investigation: LN, MGMB, IC, MB, ZZ, NR, JC

Visualization: LN, IC

Funding acquisition: JLP, MC, MGMB, LN

Project administration: MGB, JLP, MC

Supervision: LN, JLP, MC

Writing – original draft: LN, MC

Writing – review & editing: LN, MGB, IC, AB, NG, JLP, MC

## Data and materials availability

All data are available in the main text or in the supplementary materials.

