## supplemental data for "TNFRSF13B genotype governs susceptibility to ARDS through IgG-mediated complement activation"

**The PDF file includes:**

Extended Data Figs. 1-3
Extended Data Tables 1-6

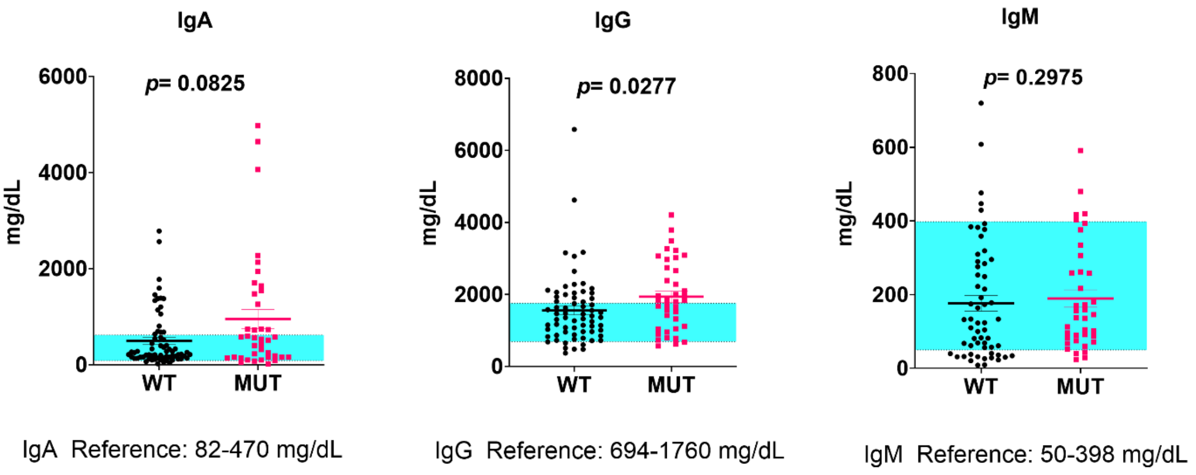


**Extended Data Fig. 1: Plasma immunoglobulin concentrations in SARS-CoV-2 infected subjects. a,** Plasma IgA, **(b)** IgG, and **(c)** IgM in *TNFRSF13B* variant carriers (MUT) compared to subjects with WT alleles in subjects infected with SARS-CoV-2 (WT, n = 67; MUT, n = 39). The blue bar depicts the normal range. Groups were compared by a two-tailed Mann-Whitney Test in GraphPad Prism. *p* values are indicated on the graphs.

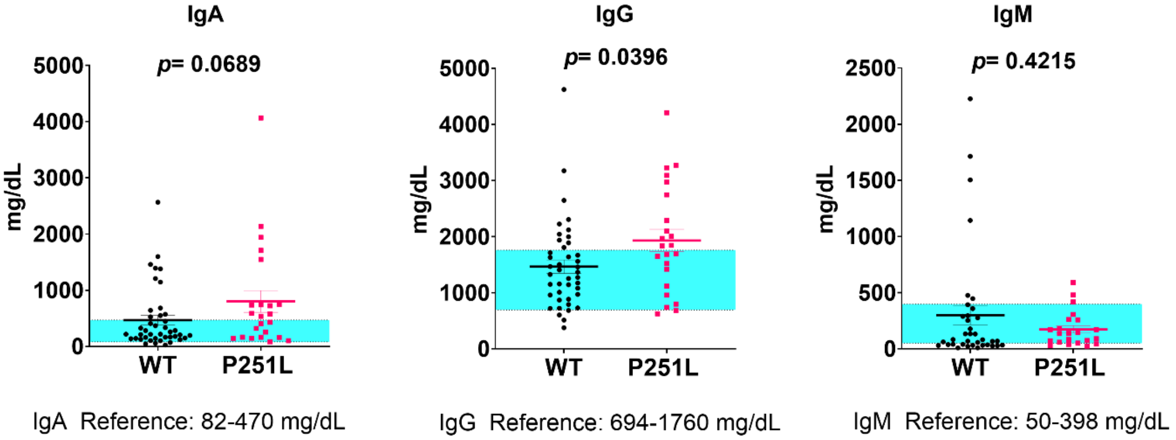


**Extended Data Fig. 2: Plasma immunoglobulin concentrations in severe SARS-CoV-2 infected subjects with WT TNFRSF13B or P251L TNFRSF13B. a,** Plasma IgA, **(b)** IgG, and **(c)** IgM in *TNFRSF13B* P251L variant carriers compared to subjects with WT alleles in the severe COVID-19 group (WT, n = 42; MUT, n = 23). The blue bar depicts the normal range. Groups were compared by a two-tailed Mann-Whitney Test in GraphPad Prism. *p* values are indicated on the graphs.

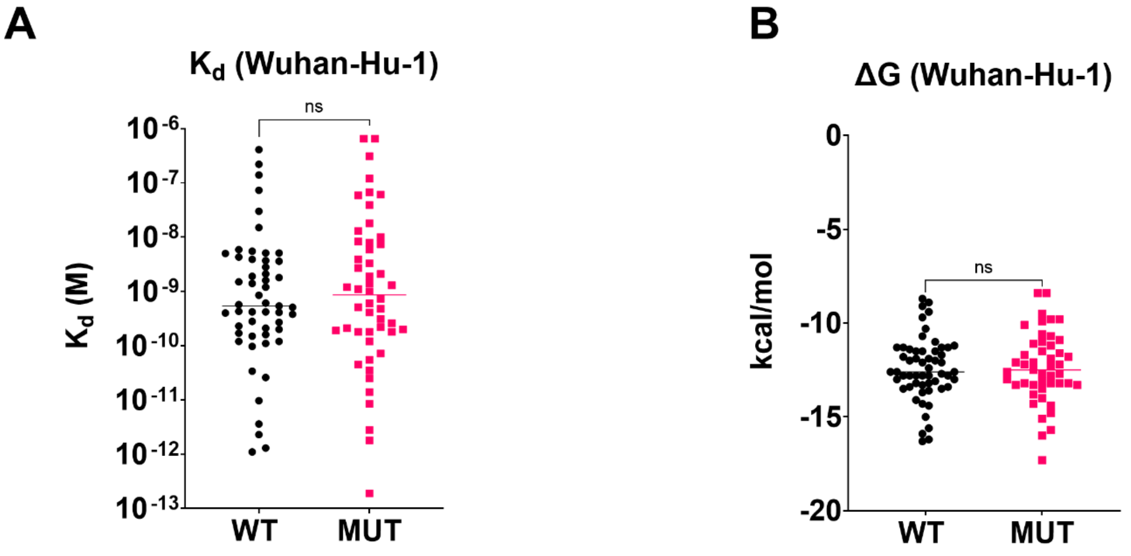


C

| | Average Contact Distance (Å) | | |
|---|---|---|---|
| Region | Higher Potency Antibodies | Lower Potency Antibodies | p-value |
| NTD | 7.2 | 7.8 | 0.7298 |
| RBD | 6.6 | 7.4 | 0.0177 |
| CTD | 8 | 7.5 | 0.2643 |
| S2 | 7.9 | 7.5 | 0.0313 |

**Higher Potency Antibodies = $IC_{50}$ of 0.01µg/mL or Less**

**Lower Potency Antibodies = $IC_{50}$ of 0.01µg/mL or More**

**Extended Data Fig. 3: *In silico* modeling of WT and *TNFRSF13B* mutant-derived antibodies bound to the SARS-CoV-2 Wuhan-Hu-1 spike protein.** Antibody heavy and light chain variable regions from WT and *TNFRSF13B* mutant (MUT) subjects were modeled using ABodyBuilder and docked onto the Wuhan-Hu-1 spike protein (PDB ID: 6VYB) with ClusPro. The analysis included 55 monoclonal antibodies from 5 WT subjects and 46 from 5 MUT subjects (1 P251L homozygous, 2 heterozygous P251L, 1 C104R, and 1 G190R, both monoallelic). **a,** Predicted binding free energy (ΔG) and **(b)** equilibrium dissociation constant ($K_d$) values were calculated using PRODIGY to compare WT and MUT antibodies. **c,** Average residue-level contact distances were calculated for the N-terminal domain (NTD), receptor-binding domain (RBD), C-terminal domain (CTD), and S2 region across antibodies classified as higher potency ($IC_{50} \leq 0.01$ µg/mL; n = 8) or lower potency ($IC_{50} \geq 0.01$ µg/mL; n = 6). Statistical comparisons for ΔG and $K_d$ were performed using Welch's t-test in GraphPad Prism. Statistical comparisons for average contact distance were performed using unpaired t-test in GraphPad Prism.

**Extended Data Table 1: Clinical and demographic characteristics of patients in the severe SARS-CoV-2 infection group.**

| Patient ID | Age | Gender | Race | TNFRSF13B Genotype | Severe Disease? | SpO2 (lowest) | Respiratory rate | Intubation | O2 flow rate (L/min) | Respiratory Symptoms/Diseases /Notes |
|---|---|---|---|---|---|---|---|---|---|---|
| UMH1 | 73 | M | White or Caucasian | P251L/WT | Yes | 94% | 16 | Yes | 4 | N/A |
| UMH2 | 72 | M | Black or African American | WT | Yes | 81% | 52 | Yes | 6 | COPD, ARDS, Pneumonia |
| UMH3 | 59 | M | Black or African American | G190R/WT | Yes | 77% | 34 | Yes | 30 | AHRF, ARDS, pneumonia |
| UMH5 | 54 | M | Black or African American | WT | Yes | 68% | 36 | Yes | 50 | ARDS, pneumonia, pulmonary embolism |
| UMH6 | 26 | M | White or Caucasian | WT | Yes | 58% | 26 | Yes | 50 | MDRO from tracheal aspirate, ARDS, Pneumonia, bacteremia, failure to liberate from the ventilator, R sided pneumothorax |
| UMH7 | 60 | M | White or Caucasian | P251L/P251L | Yes | 54% | 37 | Yes | 15 | AHRF, ARDS, pneumonia, MSSA bacteremia, shock septic |
| UMHS1 | Unknown | Unknown | Unknown | P251L/P251L | Yes | N/A | N/A | N/A | N/A | Consented at ICU/No chart access |
| UMHS2 | 37 | M | Asian | P251L/P251L | Yes | 78% | 68 | Yes | 50 | ARDS, pneumonia, pulmonary embolism |
| UMHS3 | 48 | F | White or Caucasian | V220A/WT | Yes | 87% | 18 | Yes | 6 | AHRF, ARDS, mechanically assisted ventilation |
| UMHS4 | Unknown | Unknown | Unknown | WT | Yes | N/A | N/A | N/A | N/A | Consented at ICU/No chart access |
| UMHS5 | Unknown | Unknown | Unknown | P116T/WT | Yes | N/A | N/A | N/A | N/A | Consented at ICU/No chart access |
| HFHS5 | Unknown | Unknown | Unknown | P251L/WT | Yes | N/A | N/A | N/A | N/A | pneumonia |
| COVPro-00038 | 26 | M | Black or African American | WT | Yes | 82% | 40 | Yes | 50 | pneumonia, ARDS, pulmonary embolism, ECMO |
| COVPro 00049 | 49 | M | White or Caucasian | WT | Yes | 69% | 30 | Yes | 50 | AHRF, ARDS |
| COVPro-00057 | 52 | M | White or Caucasian | P251L/P251L | Yes | 76% | 50 | Yes | 50 | ARDS, ECMO |
| COVPro-00031 | 48 | M | White or Caucasian | P251L/P251L | Yes | 90% | 22 | No | N/A | ARDS, pneumonia |
| COVPro-00078 | 45 | M | White or Caucasian | WT | Yes | 88% | N/A | Yes | 2 | N/A |
| COVPro-00041 | 55 | F | White or Caucasian | WT | Yes | 91% | 29 | Yes | 3 | pneumonia |
| COVPro-00058 | 67 | F | Black or African American | P251L/WT | Yes | <90% | 18 | Yes | 3 | pneumonia |
| COVPro-00079 | 78 | M | White or Caucasian | WT | Yes | 80% | 24 | Yes | 50 | pneumonia, ARDS, history of COPD, supplementary 02 by nasal cannula while sleeping |
| COVPro 00009 | 67 | F | Black or African American | WT | Yes | 61% | 18 | Yes | 12 | pulmonary embolism, pneumonia, ARDS |
| COVPro-00032 | 36 | M | Asian | P251L/WT | Yes | 87% | 28 | Yes | 4 | pneumonia |
| COVPro-00051 | 43 | M | Black or African American | P251L/WT | Yes | 86% | 40 | Yes | 2 | pneumonia |
| COVPro 00066 | 61 | M | White or Caucasian | WT | Yes | 91% | 20 | No | N/A | pneumonia |
| COVPro-00073 | 57 | M | Black or African American | WT | Yes | 88% | 44 | Yes | 50 | pneumonia |
| COVPro-00015 | 34 | M | Black or African American | WT | Yes | 87% | 63 | Yes | 15 | ARDS, Sepsis, pneumonia, ECMO |
| COVPro-00086 | 44 | M | White or Caucasian | WT | Yes | 84% | 32 | Yes | 15 | pneumonia, ARDS |
| COVPro-00043 | 49 | M | Asian | P251L/WT | Yes | 83% | 24 | Yes | 50 | ARDS, MDRO from tracheal aspirate |
| COVPro-00017 | 44 | M | Black or African American | WT | Yes | 85% | 26 | Yes | 50 | ARDS, pneumonia, ECMO |
| COVPro-00087 | 55 | M | Black or African American | P251L/WT | Yes | 91% | 18 | N/A | N/A | pneumonia, pulmonary embolism, Pleurisy |
| COVPro 00053 | 37 | F | Asian | P251L/P251L | Yes | 91% | N/A | N/A | N/A | pneumonia |
| COVPro-00061 | 60 | M | White or Caucasian | WT | Yes | 87% | 20 | Yes | 4 | pneumonia |
| COVPro-00013 | 58 | F | Black or African American | P251L/WT | Yes | 76% | 26 | Yes | 50 | pneumonia, ARDS, pleurisy |
| COVPro 00007 | 54 | F | White or Caucasian | V220A/WT | Yes | 88% | N/A | Yes | 15 | pneumonia, ARDS |
| COVPro-00062 | 48 | M | White or Caucasian | WT | Yes | 92% | 20 | Yes | 4 | pneumonia |
| COVPro-00023 | 57 | F | Unknown | WT | Yes | 80% | 33 | N/A | N/A | ARDS, pneumonia, ECMO, bilateral pneumotoraces |
| COVPro-00046 | 59 | M | Other | P251L/WT | Yes | 82% | 18 | N/A | N/A | pneumonia, ARDS |
| COVPro-00055 | 64 | M | White or Caucasian | P251L/WT | Yes | 84% | N/A | Yes | 10 | pneumonia, ARDS |
| COVPro-00063 | 65 | F | White or Caucasian | WT | Yes | 80% | 26 | Yes | 30 | Pneumonia |
| COVPro-00084 | 56 | M | Black or African American, White or Caucasian | WT | Yes | 87% | 40 | Yes | 6 | pneumonia, ARDS, ventilation for 2 weeks, COPD |
| COVPro 00002 | 58 | M | Black or African American | WT | Yes | 79% | 22 | Yes | 15 | ARDS, Pneumonia |
| COVPro-00048 | 51 | F | Asian | P251L/P251L | Yes | 68% | 24 | Yes | 50 | ARDS, bronchiectasis |
| COVPro-00056 | 33 | M | White or Caucasian | WT | Yes | 56% | 22 | Yes | 50 | ARDS, pneumonia |
| COVPro 00064 | 64 | M | White or Caucasian | P251L/WT | Yes | 91% | 15 | Yes | 1 | pneumonia |
| COVPro-00085 | 53 | F | Black or African American | P251L/WT | Yes | 95% | N/A | N/A | N/A | pneumonia |
| COD001-0095 | 44 | M | White or Caucasian | WT | Yes | 96% | 18 | No | N/A | Pneumonia |
| COD001-0110 | 44 | F | Black or African American | P251L/WT | Yes | 100% | 33 | Yes | N/A | Pneumonia |
| COD001-0111 | 44 | M | White or Caucasian | WT | Yes | 91% | 40 | No | N/A | Pneumonia |
| COD001-0140 | 29 | M | White or Caucasian | WT | Yes | 96% | 27 | Yes | N/A | Pneumonia |
| COD001-0147 | 50 | M | White or Caucasian | WT | Yes | 93% | 32 | No | N/A | Pneumonia |
| COD003 0005 | 45 | M | Black or African American | WT | Yes | 94% | 16 | No | N/A | Pneumonia |
| COD003-0087 | 65 | F | White or Caucasian | WT | Yes | 83% | 20 | Yes | N/A | Pneumonia, ARDS, COPD |
| COD009-0022 | 84 | F | White or Caucasian | WT | Yes | 85% | 24 | Yes | N/A | Pneumonia, ARDS, COPD |
| COD009-0024 | 42 | F | Black or African American | WT | Yes | 94% | 51 | Yes | N/A | Pneumonia |
| COD009-0027 | 38 | M | White or Caucasian | WT | Yes | 94% | 24 | No | N/A | Pneumonia |
| COD009-0030 | 85 | F | Other | WT | Yes | 93% | 26 | Yes | N/A | Pneumonia, COPD |
| COD009-0034 | 88 | M | Other | WT | Yes | 96% | 32 | No | N/A | Pneumonia |
| COD009-0037 | 72 | M | White or Caucasian | WT | Yes | 91% | 18 | Yes | N/A | Pneumonia, COPD |
| COD009-0041 | 65 | F | White or Caucasian | WT | Yes | 92% | 12 | No | N/A | Pneumonia |
| COD009-0047 | 66 | F | White or Caucasian | WT | Yes | 97% | 19 | No | N/A | Pneumonia |
| COD009 0049 | 38 | M | White or Caucasian | P251L/WT | Yes | 92% | 18 | No | N/A | Pneumonia |
| COD009-0052 | 50 | M | White or Caucasian | P251L/WT | Yes | 93% | 39 | Yes | N/A | Pneumonia, ARDS |
| COD009-0055 | 39 | F | White or Caucasian | WT | Yes | 93% | 22 | No | N/A | Pneumonia |
| COD009-0068 | 80 | F | White or Caucasian | WT | Yes | 100% | 18 | Yes | N/A | Pneumonia |
| COD009-0074 | 72 | M | White or Caucasian | WT | Yes | 93% | 27 | Yes | N/A | Pneumonia, COPD |
| COD009-0081 | 83 | F | White or Caucasian | WT | Yes | 84% | 21 | Yes | N/A | Pneumonia |

The table summarizes patient age, sex, race, and *TNFRSF13B* genotype, along with clinical parameters including $SpO_2$, lowest respiratory rate recorded during ICU admission, intubation status, oxygen flow rate, primary clinical diagnosis, and relevant physician notes.

**Extended Data Table 2: Distribution of the *TNFRSF13B* missense mutation P251L across ethnic groups in subjects with SARS-CoV-2 infection.**

| | | COVID-19 | | | | | | Severe COVID-19 | | | | | | Population | | |
|---|---|---|---|---|---|---|---|---|---|---|---|---|---|---|---|---|
| **Ethnicity** | **Missense Mutation** | **MUT** | **WT** | **Allele Frequency (%)** | **p value** | **RR** | **95% CI** | **MT** | **WT** | **Allele Frequency (%)** | **p value** | **RR** | **95% CI** | **MT** | **WT** | **Allele Frequency (%)** |
| **African American** | **P251L** | 6 | 36 | 14.29 | 0.821 | 1.075 | 0.4643 - 2.487 | 6 | 28 | 17.65 | 0.4491 | 1.382 | 0.5874 - 3.249 | 10063 | 64893 | 15.506 |
| **Asian** | **P251L** | 8 | 2 | 80.00 | **0.0003** | **13.100** | 3.148 - 54.50 | 8 | 2 | 80.00 | **0.0003** | **13.100** | 3.148 - 54.50 | 31697 | 103821 | 30.528 |
| | **P251L Hom** | 6 | 4 | 60.00 | **<0.0001** | **20.500** | 6.217 - 67.57 | 6 | 4 | 60.00 | **<0.0001** | **20.500** | 6.217 - 67.57 | 9236 | 126282 | 7.313 |
| **White or Caucasian** | **P251L** | 18 | 98 | 15.52 | 0.0893 | 1.601 | 0.9734 - 2.632 | 12 | 62 | 16.22 | 0.1374 | 1.687 | 0.9178 - 3.100 | 131542 | 1146458 | 11.474 |
| | **P251L Hom** | 8 | 108 | 6.90 | **<0.0001** | **6.778** | 3.354 - 13.69 | 6 | 68 | 8.11 | **<0.0001** | **8.074** | 3.584 - 18.18 | 13810 | 1264190 | 1.092 |

The table shows alleles frequency of WT *TNFRSF13B* (WT) or P251L *TNFRSF13B* variants (MT), *p* values, and relative risk (RR). General population allele frequencies for each ethnicity are included for comparison.

**Extended Data Table 3: Distribution of top 60 residues of anti-spike antibodies obtained from WT or P251L subjects that differ the most in contact frequency with the RBD.**

| | | WT | | | P251L | | |
|---|---|---|---|---|---|---|---|
| Residue | Motif | # of Contacts | # of Antibodies Analyzed | % of Antibody Contact | # of Contacts | # of Antibodies Analyzed | % of Antibody Contact |
| 46 | NTD | 6 | 55 | 11% | 0 | 16 | 0% |
| 69 | NTD | 0 | 55 | 0% | 2 | 16 | 13% |
| 76 | NTD | 1 | 55 | 2% | 2 | 16 | 13% |
| 97 | NTD | 1 | 55 | 2% | 2 | 16 | 13% |
| 144 | NTD | 1 | 55 | 2% | 2 | 16 | 13% |
| 148 | NTD | 1 | 55 | 2% | 2 | 16 | 13% |
| 165 | NTD | 5 | 55 | 9% | 0 | 16 | 0% |
| 182 | NTD | 1 | 55 | 2% | 2 | 16 | 13% |
| 184 | NTD | 1 | 55 | 2% | 2 | 16 | 13% |
| 252 | NTD | 1 | 55 | 2% | 2 | 16 | 13% |
| 262 | NTD | 1 | 55 | 2% | 2 | 16 | 13% |
| 355 | RBD | 0 | 55 | 0% | 2 | 16 | 13% |
| 378 | RBD | 3 | 55 | 5% | 3 | 16 | 19% |
| 380 | RBD | 5 | 55 | 9% | 3 | 16 | 19% |
| 408 | RBD | 11 | 55 | 20% | 5 | 16 | 31% |
| 416 | RBD | 4 | 55 | 7% | 3 | 16 | 19% |
| 421 | RBD | 5 | 55 | 9% | 3 | 16 | 19% |
| 424 | RBD | 5 | 55 | 9% | 5 | 16 | 31% |
| 427 | RBD | 7 | 55 | 13% | 5 | 16 | 31% |
| 449 | RBD | 5 | 55 | 9% | 4 | 16 | 25% |
| 450 | RBD | 4 | 55 | 7% | 3 | 16 | 19% |
| 455 | RBD | 1 | 55 | 2% | 2 | 16 | 13% |
| 458 | RBD | 8 | 55 | 15% | 4 | 16 | 25% |
| 466 | RBD | 4 | 55 | 7% | 3 | 16 | 19% |
| 468 | RBD | 2 | 55 | 4% | 3 | 16 | 19% |
| 473 | RBD | 6 | 55 | 11% | 4 | 16 | 25% |
| 504 | RBD | 6 | 55 | 11% | 0 | 16 | 0% |
| 551 | CTD | 7 | 55 | 13% | 0 | 16 | 0% |
| 553 | CTD | 7 | 55 | 13% | 0 | 16 | 0% |
| 554 | CTD | 6 | 55 | 11% | 0 | 16 | 0% |
| 556 | CTD | 12 | 55 | 22% | 0 | 16 | 0% |
| 568 | CTD | 7 | 55 | 13% | 0 | 16 | 0% |
| 569 | CTD | 8 | 55 | 15% | 0 | 16 | 0% |
| 588 | CTD | 9 | 55 | 16% | 0 | 16 | 0% |
| 589 | CTD | 9 | 55 | 16% | 0 | 16 | 0% |
| 591 | CTD | 6 | 55 | 11% | 0 | 16 | 0% |
| 616 | CTD | 12 | 55 | 22% | 0 | 16 | 0% |
| 617 | CTD | 6 | 55 | 11% | 0 | 16 | 0% |
| 618 | CTD | 7 | 55 | 13% | 0 | 16 | 0% |
| 644 | CTD | 10 | 55 | 18% | 0 | 16 | 0% |
| 645 | CTD | 10 | 55 | 18% | 0 | 16 | 0% |
| 646 | CTD | 13 | 55 | 24% | 0 | 16 | 0% |
| 813 | S2 | 8 | 55 | 15% | 0 | 16 | 0% |
| 815 | S2 | 6 | 55 | 11% | 0 | 16 | 0% |
| 824 | S2 | 8 | 55 | 15% | 0 | 16 | 0% |
| 828 | S2 | 11 | 55 | 20% | 0 | 16 | 0% |
| 855 | S2 | 13 | 55 | 24% | 0 | 16 | 0% |
| 860 | S2 | 6 | 55 | 11% | 0 | 16 | 0% |
| 866 | S2 | 6 | 55 | 11% | 0 | 16 | 0% |
| 867 | S2 | 12 | 55 | 22% | 0 | 16 | 0% |
| 868 | S2 | 8 | 55 | 15% | 0 | 16 | 0% |
| 1084 | S2 | 13 | 55 | 24% | 2 | 16 | 13% |
| 1086 | S2 | 13 | 55 | 24% | 2 | 16 | 13% |
| 1123 | S2 | 6 | 55 | 11% | 0 | 16 | 0% |
| 1125 | S2 | 9 | 55 | 16% | 1 | 16 | 6% |
| 1136 | S2 | 12 | 55 | 22% | 2 | 16 | 13% |

Top 60 residues showing the largest absolute differences in antibody–spike contact frequency between WT and P251L antibodies, with contact frequencies shown as percentages for each group.

**Extended Data Table 4: Distribution of top 60 residues of anti-spike antibodies obtained from WT or C104R subjects that differ the most in contact frequency with the RBD**

| | | WT | | | C104R | | |
|---|---|---|---|---|---|---|---|
| Residue | Motif | # of Contacts | # of Antibodies Analyzed | % of Antibody Contact | # of Contacts | # of Antibodies Analyzed | % of Antibody Contact |
| 32 | NTD | 1 | 55 | 2% | 2 | 9 | 22% |
| 33 | NTD | 2 | 55 | 4% | 2 | 9 | 22% |
| 58 | NTD | 0 | 55 | 0% | 2 | 9 | 22% |
| 59 | NTD | 2 | 55 | 4% | 2 | 9 | 22% |
| 164 | NTD | 7 | 55 | 13% | 0 | 9 | 0% |
| 215 | NTD | 1 | 55 | 2% | 2 | 9 | 22% |
| 216 | NTD | 2 | 55 | 4% | 2 | 9 | 22% |
| 218 | NTD | 2 | 55 | 4% | 3 | 9 | 33% |
| 219 | NTD | 2 | 55 | 4% | 2 | 9 | 22% |
| 220 | NTD | 2 | 55 | 4% | 2 | 9 | 22% |
| 278 | NTD | 2 | 55 | 4% | 2 | 9 | 22% |
| 280 | NTD | 1 | 55 | 2% | 2 | 9 | 22% |
| 281 | NTD | 3 | 55 | 5% | 2 | 9 | 22% |
| 286 | NTD | 2 | 55 | 4% | 2 | 9 | 22% |
| 346 | RBD | 7 | 55 | 13% | 0 | 9 | 0% |
| 378 | RBD | 3 | 55 | 5% | 2 | 9 | 22% |
| 403 | RBD | 3 | 55 | 5% | 3 | 9 | 33% |
| 405 | RBD | 6 | 55 | 11% | 3 | 9 | 33% |
| 414 | RBD | 11 | 55 | 20% | 3 | 9 | 33% |
| 415 | RBD | 11 | 55 | 20% | 3 | 9 | 33% |
| 421 | RBD | 5 | 55 | 9% | 2 | 9 | 22% |
| 446 | RBD | 7 | 55 | 13% | 0 | 9 | 0% |
| 458 | RBD | 8 | 55 | 15% | 4 | 9 | 44% |
| 460 | RBD | 9 | 55 | 16% | 3 | 9 | 33% |
| 473 | RBD | 6 | 55 | 11% | 3 | 9 | 33% |
| 474 | RBD | 0 | 55 | 0% | 2 | 9 | 22% |
| 476 | RBD | 5 | 55 | 9% | 2 | 9 | 22% |
| 489 | RBD | 2 | 55 | 4% | 2 | 9 | 22% |
| 505 | RBD | 5 | 55 | 9% | 2 | 9 | 22% |
| 551 | CTD | 7 | 55 | 13% | 0 | 9 | 0% |
| 553 | CTD | 7 | 55 | 13% | 0 | 9 | 0% |
| 556 | CTD | 12 | 55 | 22% | 0 | 9 | 0% |
| 568 | CTD | 7 | 55 | 13% | 0 | 9 | 0% |
| 569 | CTD | 8 | 55 | 15% | 0 | 9 | 0% |
| 588 | CTD | 9 | 55 | 16% | 0 | 9 | 0% |
| 589 | CTD | 9 | 55 | 16% | 0 | 9 | 0% |
| 602 | CTD | 1 | 55 | 2% | 2 | 9 | 22% |
| 603 | CTD | 2 | 55 | 4% | 3 | 9 | 33% |
| 604 | CTD | 1 | 55 | 2% | 2 | 9 | 22% |
| 605 | CTD | 1 | 55 | 2% | 2 | 9 | 22% |
| 606 | CTD | 1 | 55 | 2% | 2 | 9 | 22% |
| 607 | CTD | 1 | 55 | 2% | 2 | 9 | 22% |
| 616 | CTD | 12 | 55 | 22% | 0 | 9 | 0% |
| 618 | CTD | 7 | 55 | 13% | 0 | 9 | 0% |
| 644 | CTD | 10 | 55 | 18% | 0 | 9 | 0% |
| 645 | CTD | 10 | 55 | 18% | 0 | 9 | 0% |
| 646 | CTD | 13 | 55 | 24% | 0 | 9 | 0% |
| 674 | CTD | 1 | 55 | 2% | 2 | 9 | 22% |
| 813 | S2 | 8 | 55 | 15% | 0 | 9 | 0% |
| 824 | S2 | 8 | 55 | 15% | 0 | 9 | 0% |
| 828 | S2 | 11 | 55 | 20% | 0 | 9 | 0% |
| 855 | S2 | 13 | 55 | 24% | 0 | 9 | 0% |
| 867 | S2 | 12 | 55 | 22% | 0 | 9 | 0% |
| 868 | S2 | 8 | 55 | 15% | 0 | 9 | 0% |
| 1083 | S2 | 7 | 55 | 13% | 0 | 9 | 0% |
| 1084 | S2 | 13 | 55 | 24% | 1 | 9 | 11% |

Top 60 residues showing the largest absolute differences in antibody–spike contact frequency between WT and C104R antibodies, with contact frequencies shown as percentages for each group.

**Extended Data Table 5: Distribution of top 60 residues of anti-spike antibodies obtained from WT or G190R subjects that differ the most in contact frequency with the RBD**

| | | WT | | | G190R | | |
|---|---|---|---|---|---|---|---|
| Residue | Motif | # of Contacts | # of Antibodies Analyzed | % of Antibody Contact | # of Contacts | # of Antibodies Analyzed | % of Antibody Contact |
| 46 | NTD | 6 | 55 | 11% | 0 | 21 | 0% |
| 153 | NTD | 5 | 55 | 9% | 0 | 21 | 0% |
| 170 | NTD | 1 | 55 | 2% | 2 | 21 | 10% |
| 173 | NTD | 1 | 55 | 2% | 2 | 21 | 10% |
| 174 | NTD | 1 | 55 | 2% | 2 | 21 | 10% |
| 175 | NTD | 0 | 55 | 0% | 2 | 21 | 10% |
| 176 | NTD | 0 | 55 | 0% | 2 | 21 | 10% |
| 177 | NTD | 0 | 55 | 0% | 2 | 21 | 10% |
| 207 | NTD | 0 | 55 | 0% | 2 | 21 | 10% |
| 217 | NTD | 0 | 55 | 0% | 2 | 21 | 10% |
| 297 | NTD | 0 | 55 | 0% | 2 | 21 | 10% |
| 300 | NTD | 0 | 55 | 0% | 2 | 21 | 10% |
| 345 | RBD | 5 | 55 | 9% | 0 | 21 | 0% |
| 346 | RBD | 7 | 55 | 13% | 0 | 21 | 0% |
| 408 | RBD | 11 | 55 | 20% | 2 | 21 | 10% |
| 409 | RBD | 8 | 55 | 15% | 1 | 21 | 5% |
| 445 | RBD | 5 | 55 | 9% | 0 | 21 | 0% |
| 446 | RBD | 7 | 55 | 13% | 1 | 21 | 5% |
| 455 | RBD | 1 | 55 | 2% | 3 | 21 | 14% |
| 456 | RBD | 3 | 55 | 5% | 3 | 21 | 14% |
| 473 | RBD | 6 | 55 | 11% | 4 | 21 | 19% |
| 475 | RBD | 6 | 55 | 11% | 4 | 21 | 19% |
| 489 | RBD | 2 | 55 | 4% | 3 | 21 | 14% |
| 501 | RBD | 5 | 55 | 9% | 0 | 21 | 0% |
| 503 | RBD | 5 | 55 | 9% | 0 | 21 | 0% |
| 504 | RBD | 6 | 55 | 11% | 0 | 21 | 0% |
| 561 | CTD | 0 | 55 | 0% | 2 | 21 | 10% |
| 581 | CTD | 0 | 55 | 0% | 2 | 21 | 10% |
| 582 | CTD | 0 | 55 | 0% | 2 | 21 | 10% |
| 584 | CTD | 0 | 55 | 0% | 2 | 21 | 10% |
| 588 | CTD | 9 | 55 | 16% | 0 | 21 | 0% |
| 589 | CTD | 9 | 55 | 16% | 0 | 21 | 0% |
| 616 | CTD | 12 | 55 | 22% | 2 | 21 | 10% |
| 617 | CTD | 6 | 55 | 11% | 0 | 21 | 0% |
| 618 | CTD | 7 | 55 | 13% | 1 | 21 | 5% |
| 644 | CTD | 10 | 55 | 18% | 2 | 21 | 10% |
| 645 | CTD | 10 | 55 | 18% | 1 | 21 | 5% |
| 646 | CTD | 13 | 55 | 24% | 2 | 21 | 10% |
| 654 | CTD | 0 | 55 | 0% | 2 | 21 | 10% |
| 691 | S2 | 0 | 55 | 0% | 2 | 21 | 10% |
| 692 | S2 | 0 | 55 | 0% | 2 | 21 | 10% |
| 814 | S2 | 5 | 55 | 9% | 0 | 21 | 0% |
| 815 | S2 | 6 | 55 | 11% | 0 | 21 | 0% |
| 828 | S2 | 11 | 55 | 20% | 2 | 21 | 10% |
| 855 | S2 | 13 | 55 | 24% | 0 | 21 | 0% |
| 860 | S2 | 6 | 55 | 11% | 0 | 21 | 0% |
| 867 | S2 | 12 | 55 | 22% | 2 | 21 | 10% |
| 1084 | S2 | 13 | 55 | 24% | 7 | 21 | 33% |
| 1085 | S2 | 5 | 55 | 9% | 4 | 21 | 19% |
| 1098 | S2 | 8 | 55 | 15% | 1 | 21 | 5% |
| 1101 | S2 | 11 | 55 | 20% | 2 | 21 | 10% |
| 1103 | S2 | 8 | 55 | 15% | 0 | 21 | 0% |
| 1112 | S2 | 4 | 55 | 7% | 4 | 21 | 19% |
| 1122 | S2 | 3 | 55 | 5% | 4 | 21 | 19% |
| 1123 | S2 | 6 | 55 | 11% | 4 | 21 | 19% |
| 1127 | S2 | 7 | 55 | 13% | 5 | 21 | 24% |
| 1136 | S2 | 12 | 55 | 22% | 2 | 21 | 10% |
| 1138 | S2 | 13 | 55 | 24% | 7 | 21 | 33% |
| 1139 | S2 | 7 | 55 | 13% | 1 | 21 | 5% |
| 1143 | S2 | 11 | 55 | 20% | 6 | 21 | 29% |

Top 60 residues showing the largest absolute differences in antibody–spike contact frequency between WT and G190R antibodies, with contact frequencies shown as percentages for each group.

**Extended Data Table 6: Multivariable linear regression analysis of IgG sialylation and galactosylation.**

### IgG Sialylation

| Variable | β coefficient | Standard Error | t | *p value* |
|---|---|---|---|---|
| *TNFRSF13B Variant* | -2.84 | 1.44 | -1.981 | 0.051 |
| *Age* | 0.013 | 0.059 | 0.218 | 0.828 |
| *Male Sex* | 1.17 | 1.51 | 0.775 | 0.442 |
| *BMI* | -0.154 | 0.084 | -1.82 | 0.075 |
| *Comorbidity* | -1.62 | 1.62 | -1.00 | 0.319 |
| *Days from COVID-19 diagnosis to sample collection* | -0.018 | 0.0192 | -0.97 | 0.335 |

### IgG Galactosylation

| Variable | β coefficient | Standard Error | t | *p value* |
|---|---|---|---|---|
| *TNFRSF13B Variant* | -4.53 | 1.32 | -3.44 | 0.001 |
| *Age* | -0.012 | 0.055 | -0.23 | 0.823 |
| *Male Sex* | 2.35 | 1.39 | 1.69 | 0.097 |
| *BMI* | -0.173 | 0.078 | -2.23 | 0.031 |
| *Comorbidity* | -2.63 | 1.48 | -1.77 | 0.083 |
| *Days from COVID-19 diagnosis to sample collection* | 0.005 | 0.018 | 0.31 | 0.758 |

Multivariable linear regression models were used to assess the association between TNFRSF13B variant status, IgG sialylation, and IgG galactosylation, while adjusting for potential demographic and clinical covariates. Independent variables included TNFRSF13B genotype (WT vs variant), age, sex, body mass index (BMI), comorbidity status, and days from COVID-19 diagnosis to plasma collection. β represents the regression coefficient indicating the estimated change in glycosylation score associated with each predictor variable while holding other variables constant. t represents the t-statistic used to test whether the coefficient differs from zero. *p* values correspond to two-sided tests of statistical significance for each coefficient. Analyses included 62 total subjects (WT, n = 40; TNFRSF13B variant, n = 22).
